# Patterns and drivers of 43,617 mosaic chromosomal alterations in blood

**DOI:** 10.1101/2025.07.30.25332451

**Authors:** David Tang, Nolan Kamitaki, Ronen E. Mukamel, Simone Rubinacci, Po-Ru Loh

## Abstract

Clonal expansions of hematopoietic cells carrying mosaic chromosomal alterations (mCAs) are commonly detectable in elderly individuals. Here, we studied 43,617 autosomal mCAs that we ascertained in 484,081 UK Biobank participants using new, high-resolution computational methods to analyze blood-derived whole-genome sequencing data. Shorter mCAs (≤1 Mb) clustered at 53 genomic hotspots (34 previously undetected), several of which implicated chromosomal fragile sites as a recurrent source of somatic deletions. Chronic lymphocytic leukemia (CLL)-associated deletions at 13q14 were detectable in 1% of 65- to 70-year-old individuals—a five-fold higher rate than previously observed—suggesting opportunities for incorporating this mosaic mutation in clinical screening and in genetic association studies. Rare protein-coding variants in 38 genes associated (*p*<1.2×10^-5^; FDR<0.01) with clonal expansions of copy-neutral loss of heterozygosity (CN-LOH) mutations that modified the allelic dosages of these variants, implicating genes in DNA damage response, cell cycle, cytokine signaling, and protein ubiquitination pathways as targets of clonal selection via CN-LOH-induced allelic substitution. For several genes with proliferation-increasing functions, CN-LOH mutations produced revertant mosaicism, removing deleterious inherited variants. Polygenic effects of common inherited variants within CN-LOH mutations also contributed modestly to clonal selection. These results show that our blood genomes are constantly evolving as we age and often accrue mCAs in predictable ways.

## Introduction

Hematopoietic stem and progenitor cells typically acquire 500 to 1,500 somatic mutations across an individual’s lifespan^1–3^. While most somatic mutations do not alter a cell’s clonal fitness, some mutations confer a proliferative advantage to the cell and its progeny, such that they clonally expand to constitute an outsized proportion of the individual’s blood cells^3–5^. This phenomenon, termed clonal hematopoiesis (CH), commonly occurs during aging, such that expansions with several-percent clonal fractions are frequently observed in elderly individuals^6–9^, and subtler clonal expansions are ubiquitous by middle age^10^. CH confers tenfold increased risk of progression to hematologic malignancies^6–9^ and is also a risk factor for several other adverse health outcomes^11–13^.

Clonal hematopoiesis can be classified by the types of mutations carried in expanded clones: clonal hematopoiesis of indeterminate potential (CHIP) involves somatic SNVs and indels in leukemia driver genes^8,9^, while CH with mosaic chromosomal alterations (mCAs) involves megabase-to chromosome-scale mutations^6,7^. Most hematopoietic clones have unknown drivers (CH-UD)^3,8,14,15^. Inherited genetic variation modulates risk of all types of CH, exhibiting some shared but some distinct effects on CHIP^16–20^, autosomal^20–23^ and sex chromosome mCAs^24,25^, and CH-UD^14,15^. Similarly, different CH mutations increase risk of different blood cancers^26,27^ and other diseases^13^. Here, we focus on autosomal mCAs, which are more readily detectable at lower cell fractions compared to point mutations^21–23,28,29^, include common drivers of lymphoid malignancies^26,30,31^, and provide unique insight into genetic drivers of CH by their action on inherited and acquired variation^21–23,32,33^.

Despite remarkable progress over the past decade in understanding the causes and consequences of CH^13^, many questions remain, particularly regarding mCAs. Nearly all large-scale studies of mCAs to date have ascertained mCAs from SNP-array allelic intensity data^6,7,12,21–26,28^, limiting detection of shorter mCAs (∼1Mb) and mCAs with low clonal fractions (<1%). Additionally, the sample sizes of existing autosomal mCA call sets have not yet permitted precise natural history studies of specific mCAs. Recent work has demonstrated that analyzing whole-genome sequencing (WGS) data can increase mCA detection sensitivity^29,34^. Population sequencing data sets also provide the opportunity to more comprehensively explore the influences of inherited rare variants on clonal expansion of mCAs, building on results obtained to date^20,22,29^. Here, we developed a new computational method optimized for detecting mCAs from WGS data and applied it to UK Biobank^35,36^, generating a rich catalog of mCAs and uncovering new insights into their genetic drivers and progression to malignancy.

### Analysis of UK Biobank WGS data detects 43,617 autosomal mCAs

Our new method to detect mCAs from high-coverage (∼30x) WGS data improves upon the MoChA software pipeline^21,22^ by: (1) carefully modeling WGS depth-of-coverage and conservatively filtering kilobase-scale germline CNVs; (2) counting sequenced DNA fragments derived from each of an individual’s two haplotypes; and (3) calling mCAs by analyzing allelic imbalance and determining copy-number states from denoised read-depth data (**Fig. 1a**, **Supplementary Fig. 1, and Methods**).

**Figure 1:**
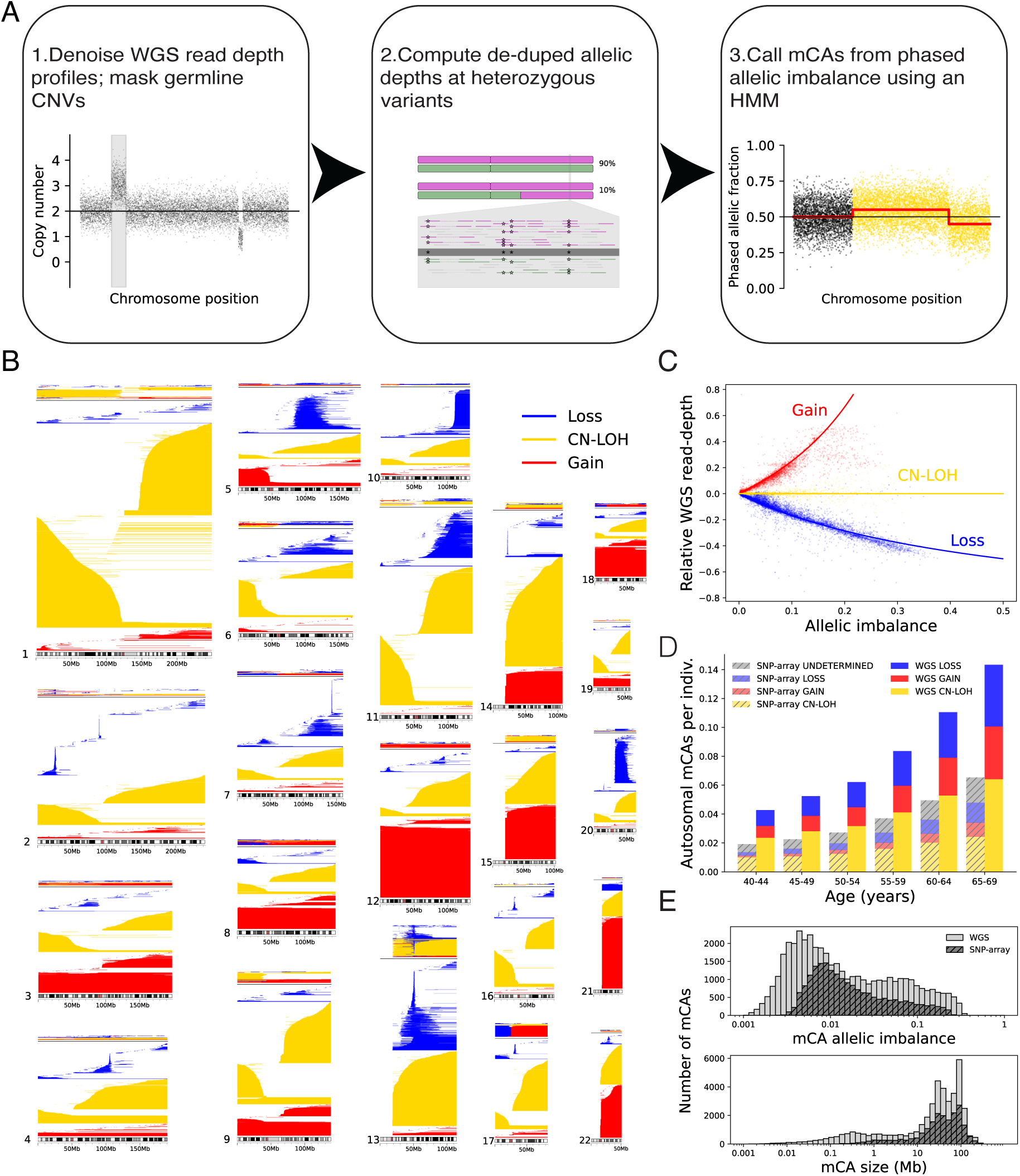
Detection of 43,617 autosomal mCAs from WGS of 484,081 UKB participants. **(A)** Overview of the computational pipeline for calling mCAs from WGS data. **(B)** Genomic “pileup” plot showing chromosomal coordinates of mCA calls. Each y-coordinate above a chromosome corresponds to a single individual and shows all mCAs called on that chromosome. Individuals with multiple mCA calls on the chromosome are shown on top, followed by individuals with only a single mosaic loss (blue), CN-LOH (gold), or gain (red) on the chromosome. **(C)** WGS read-depth deviation (i.e., fractional increase or decrease relative to expectation assuming diploid copy number) and allelic imbalance within the boundaries of mCAs cluster along three curves corresponding to the relationship between these quantities for duplications (gain), CN-LOH mutations, and deletions (loss). Each mCA was assigned a copy-number state based on linear separators, with mCAs in V(D)J recombination regions treated separately (**Methods**). **(D)** WGS-based mCA analysis detected twice as many mCAs as SNP-array-based analysis across all age groups in UKB. **(E)** WGS-based mCA calls include many more mCAs with lower clonal fractions and many more short mCAs compared to SNP-array-based calls.

Applying this approach to blood-derived WGS data for 484,081 UKB participants (generally 40–70 years old at sample acquisition) detected 43,617 autosomal mCAs in 35,033 individuals (**Fig. 1b,c** and **Supplementary Tables 1** and **2**). This represented a 2-fold increase in detection sensitivity compared to our previous SNP-array-based analysis of the same cohort^22^ (**Fig. 1d**). Beyond improving sensitivity, the WGS-based pipeline assigned copy-number state to all mCAs (**Fig. 1c** and **Supplementary Fig. 2a**), unlike SNP-array analysis (**Fig. 1d**), and mCA calls required minimal filtering for technical artifacts (**Methods**), unlike previous analyses of SNP-array data^21,22^ and WGS data^29^.

Multiple lines of evidence supported the validity of the mCA calls. First, the genomic distributions of loss, copy-neutral loss of heterozygosity (CN-LOH), and gain mCAs (**Fig. 1b**) were broadly consistent with previous studies^6,7,22,23,28,29^. Second, mCA detections increased markedly with age, from 0.04 autosomal mCAs per individual aged 40–44 years to 0.14 mCAs per individual aged 65–69 years (**Fig. 1d**). Third, whole-exome sequencing (WES) data from UKB^37^ replicated the allelic imbalances detected from WGS at expected rates (**Supplementary Fig. 2b**). Fourth, the WGS call set largely comprised a superset of the SNP-array-based mCA calls^22^: among 16,978 individuals with an mCA called from SNP-array data, 15,383 (91%) had an mCA called from WGS.

WGS-based mCA analysis particularly improved detection sensitivity for two classes of mCAs: (i) large mCAs with low clonal fractions and (ii) small mCAs with moderate-to-high clonal fractions (**Fig. 1e**). Specifically, the WGS analysis detected 2.8-fold more mCAs with allelic imbalance <0.01 (i.e., <51% representation of the overrepresented haplotype) and 10.5-fold more mCAs shorter than 1 Mb (**Fig. 1e**). The WGS analysis also identified 5.6-fold more whole chromosome CN-LOH mutations (624 versus 111 from SNP-array analysis), reflecting the low clonal fractions of most somatic isodisomies.

### WGS-based analysis identifies focal mCAs and mCA breakpoints

The increased power to detect short, interstitial mCAs from WGS data enabled a deeper characterization of “hotspots” at which short mCAs are recurrently found in blood DNA. Several hotspots of recurrent somatic deletions have previously been observed, particularly at *DLEU2* (13q14), *DNMT3A*, and *TET2*^7,21,28^. Here, higher-resolution WGS-based analysis identified 41 genomic loci (34 new relative to SNP-array analysis^22^) at which at least 10 UKB participants carried sub-megabase somatic deletions, along with 12 sites of recurrent short duplications (**Fig. 2a,b** and **Supplementary Table 3**). Duplication hotspots have not been observed from SNP-array data, in which short mosaic duplications are typically undetectable. To validate these new interstitial mCA hotspots, we searched for WGS read pairs that mapped “discordantly” to opposite ends of an mCA (within 50kb of the breakpoints) with strand orientations consistent with the putative mosaic mutation. Such read pairs are sometimes generated when a sequenced DNA fragment spans an mCA breakpoint (in a cell carrying the mutation) and are rarely observed by chance. For 48 of the 53 recurrent short mCAs, we observed discordant read support for >25% of mCA calls (**Supplementary Table 3**). The mCA calls at each hotspot exhibited varying breakpoint locations and varying levels of allelic imbalance, further supporting their somatic origin (**Fig. 2a** and **Supplementary Table 3**).

**Figure 2:**
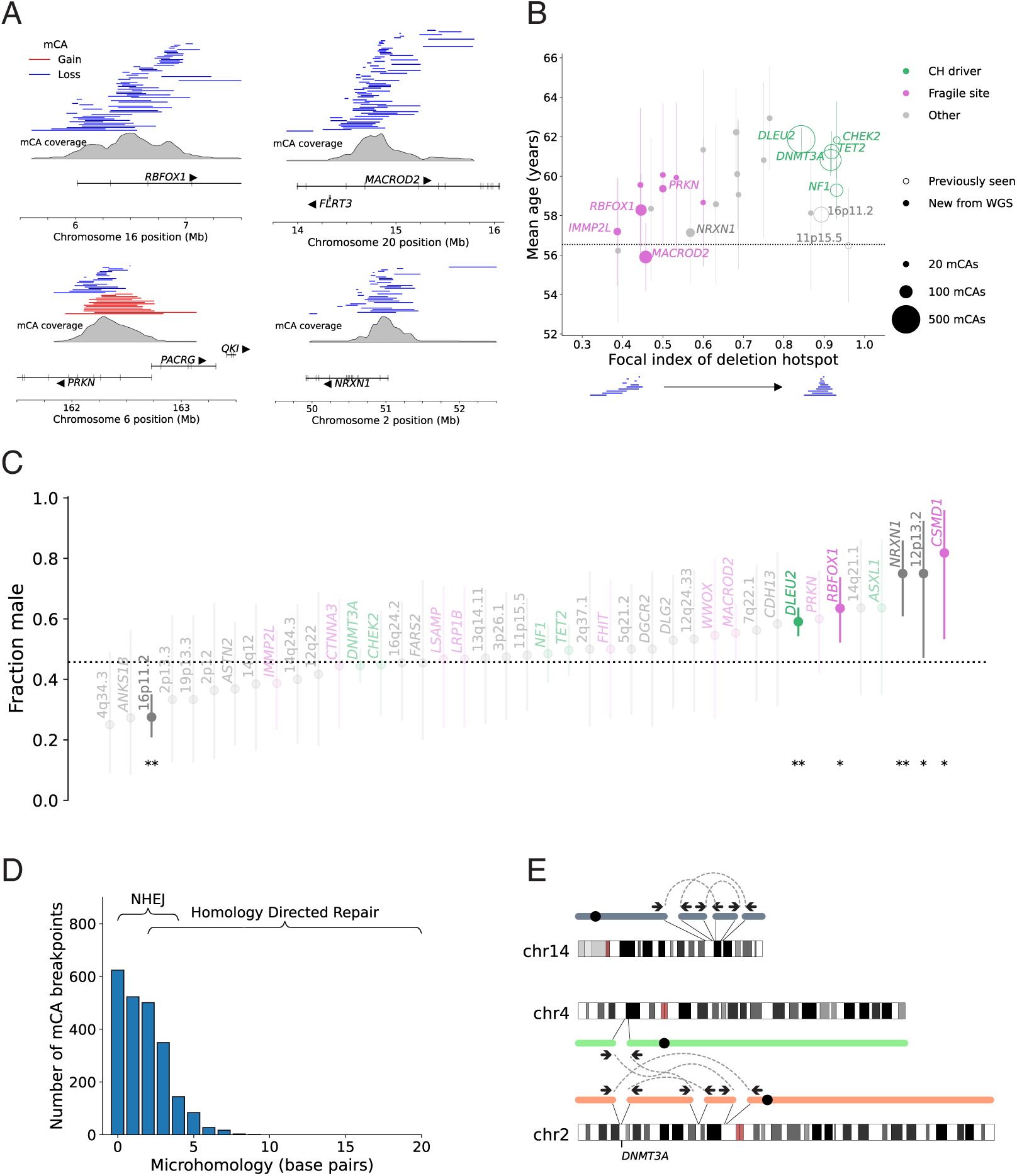
Hotspots of focal mosaic deletions and analysis of mCA breakpoints. **(A)** Pileups of mCAs at four loci where focal deletions were newly detected from WGS-based analysis. Approximate breakpoints were determined from discordant read pairs and are accurate to ∼1kb. **(B)** Properties of focal deletions observed in at least 10 UKB participants. The focal index of a deletion hotspot is a measure of the extent to which deletions at the locus overlap a single shared deletion region (**Methods**). Deletions at fragile sites (pink) tended to be less age-enriched and less focal compared to deletions at clonal hematopoiesis (CH) driver genes (green). Deletion hotspots indicated as “new from WGS” (filled circles) are those that were detected at least 5 times more frequently from WGS data compared to SNP-array data. Error bars, 95% CIs. **(C)** Fraction of carriers of each of the 41 focal deletions who are male. Error bars, 95% CIs. * *P* < 0.05; ** *P* < 0.05, Bonferroni-adjusted. **(D)** Distribution of the number of base pairs of microhomology between the left and right breakpoints of mosaic deletions for which split reads resolved the breakpoints to base-pair resolution. **(E)** Two examples of mosaic complex SVs detected in the blood of healthy individuals by analyzing WGS read pairs. In the second example, the set of rearrangements and translocations include a deletion of *DNMT3A*, which presumably contributed to the clonal expansion.

Several newly-identified deletion hotspots exhibited characteristics different from previously observed focal deletions, suggesting distinct etiology. Whereas deletions at previously-identified hotspots tended to vary in length but consistently overlap a single target gene (e.g., *DNMT3A*, *TET2*, *NF1*, *CHEK2*), deletions at the new loci tended to be shorter and less focal, typically spanning 100–700 kb with breakpoints distributed throughout the locus and no consensus deleted region (**Fig. 2a,b** and **Supplementary Table 3**). These less-focal deletions tended to be less age-enriched and to overlap fragile sites commonly deleted in cancers (e.g., *MACROD2* and *RBFOX1*)^38^ (**Fig. 2b** and **Supplementary Table 3**), suggesting that some of the deletions might have originated from mutations at fragile sites early in development rather than rising in frequency due to clonal selection during aging. Among 19 autosomal fragile sites previously observed in cancer genomes^38^, 10 fragile sites were mCA hotspots in blood. Three focal deletions exhibited Bonferroni-significant sex biases (*P*<0.05/41) relative to the cohort average (47% male): del(16p11.2) was more frequent in women^21,22^, while deletions at *DLEU2* and *NRXN1* were more frequent in men (**Fig. 2c** and **Supplementary Table 3**). Mosaic deletions at *NRXN1* observed in blood have previously been implicated in schizophrenia^39^.

To learn about the DNA repair mechanisms that give rise to interstitial deletions in blood DNA, we analyzed read-level data to resolve the breakpoints of a subset of mCAs. Discordant read pairs localized the breakpoints of 3,500 interstitial deletions (28% of deletions, including 48% of those with cell fraction >10%) to a resolution of ∼1kb, and chimeric (“split”) reads fully resolved 2,274 deletions at base-pair resolution. The majority of fully-resolved deletions (88%) had little or no microhomology at their breakpoints (at most 3bp, with 0bp being most common; **Fig. 2d**), suggesting that non-homologous end joining (NHEJ) is the double-strand break repair mechanism^40^ responsible for most mosaic deletions in blood. Other deletions exhibited intermediate amounts of microhomology (usually up to 8bp; **Fig. 2d**), indicating that microhomology mediated repair mechanisms also contribute to mCA formation.

Whole-genome sequencing data also provided the opportunity to identify several complex structural variants (SVs) in the blood of apparently healthy, cancer-free individuals: 159 individuals had discordant read pairs linking mCAs called on separate chromosomes to the same mutational event, and we fully reconstructed complex mosaic SVs for 22 individuals, revealing translocations, inversions, and complex rearrangements (**Fig. 2e**).

### Clonal expansions of CLL-associated 13q14 deletions commonly arise during aging

Our new mCA call set contained thousands of chromosomal alterations often seen in chronic lymphocytic leukemia (CLL), enabling deeper study of these mCAs and their progression to CLL. Deletion of 13q14 and trisomy 12 are the two most common mCAs in CLL (with most CLL genomes containing at least one of these two mCAs^30^), and our previous SNP-array analysis detected each alteration in 0.1% of UKB participants, conferring 100- to 200-fold increased risk of incident CLL^22^. Here, WGS-based analysis detected twice as many mCAs of each type (1,141 deletions of 13q14 and 929 trisomy 12 events; **Fig. 3a**). However, given that many 13q14 deletions are short deletions of a focal 1Mb region (**Fig. 1b**), we reasoned that a targeted analysis of WGS read-depth in this region might uncover even more mosaic 13q14 deletions present at low cell fractions. This approach extended the lower limit of detectability to a cell fraction of 2% and increased the number of detected 13q14 deletions to 2,881—a five-fold increase over the SNP-array analysis—with prevalence rising with age from 0.1% at age 40 to 1.1% by age 70 (**Fig. 3a**, **Supplementary Fig. 3**, and **Supplementary Table 4**).

**Figure 3:**
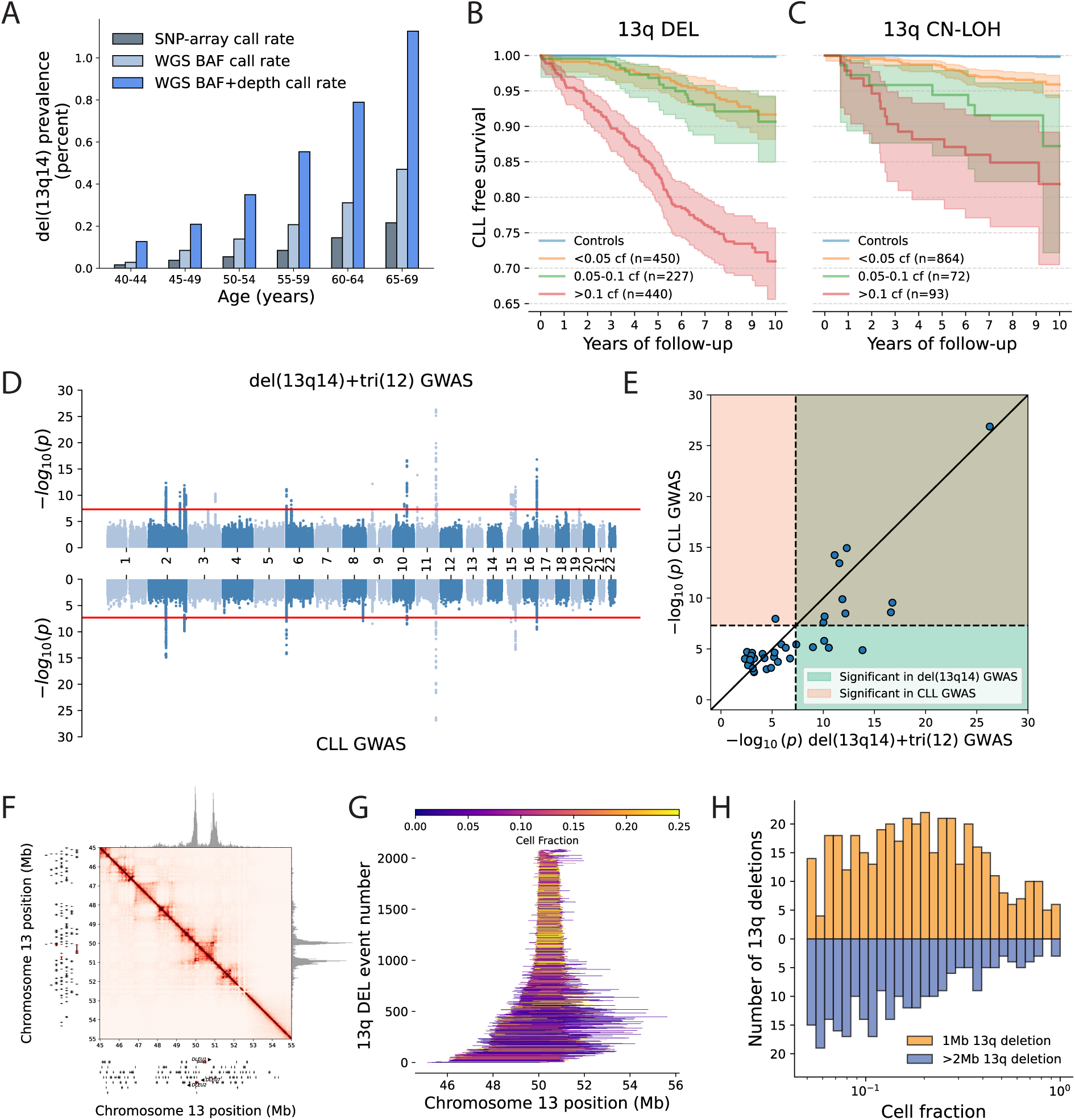
Causes and consequences of mosaic 13q14 deletion from 2,881 cases in UKB. **(A)** Prevalence of mosaic 13q14 deletions detected in blood samples from UKB participants by analyzing either SNP-array allelic imbalance data, WGS allelic imbalance data, or WGS allelic imbalance and WGS read-depth data (union of calls from the two WGS-based approaches). **(B)** Kaplan-Meier survival curves for rates of progression to chronic lymphocytic leukemia (CLL) for individuals with del(13q14) clones of various sizes (cf, cell fraction; shaded regions, 95% CIs). Individuals with 13q CN-LOH calls were excluded, and del(13q14) clones were restricted to those with breakpoints localized by discordant read pairs (allowing more accurate estimation of clonal fraction; **Methods**). Analyses were restricted to European-ancestry individuals without blood cancer diagnoses at assessment, and controls were age- and sex-matched. **(C)** Analogous CLL-free survival curves for individuals with 13q CN-LOH clones of various sizes. **(D)** Comparison of GWAS on WGS-derived del(13q14) or trisomy 12 status versus CLL case status in UKB (including up to 15 years of incident cases). **(E)** Comparison of *p*-values from the del(13q14)+tri(12) GWAS (x-axis) and the CLL GWAS (y-axis) at CLL-associated loci from Law et al. 2017^45^. For each index variant from Law et al. 2017, the plotted *p*-values indicate the most significant associations observed within 500kb in each GWAS in UKB. Dashed lines and shading correspond to the standard genome wide significance threshold of 5×10^-8^. **(F)** Hi-C contacts in naïve B-cells^46^ suggest frequent physical proximity between the most common left and right breakpoints of 13q14 deletions. **(G)** Pileups of 2,087 mosaic 13q14 deletions with breakpoints localized by discordant read pairs in 1,590 individuals with 13q14 deletions (see **Methods**). Deletions are colored by estimated cell fraction. **(H)** The cell fraction distributions of long versus short 13q14 deletions (restricted to deletions with cell fraction >5%) show that short ∼1Mb deletions (0.9–1.1Mb) tend to rise to high cell fraction more often than longer deletions (>2Mb).

Most 13q CN-LOH events, which are also associated with strongly increased CLL risk^22^, appeared to act on 13q14 deletions. Of the 199 individuals with a high-cell-fraction (>5%) CN-LOH mutation on 13q, 119 had evidence of a 13q14 deletion made biallelic by the CN-LOH (**Supplementary Fig. 4a**). However, a substantial minority (38%) had no evidence of reduced copy number at 13q14 (relative read-depth >0.99; **Supplementary Fig. 4a**), suggesting that 13q CN-LOH mutations—nearly all of which overlap 13q14 (**Fig. 1b**)—also commonly act on other genetic or epigenetic modifications that inhibit tumor suppression function at 13q14^41^.

To study the natural history of progression from 13q mCAs to CLL, we analyzed cancer outcome data accrued across up to 15 years of follow-up. Deletions of 13q14 that were likely to be monoallelic (based on the absence of 13q CN-LOH) progressed to CLL at rates that increased with clone size: 10-year CLL-free survival rates (excluding deaths from other causes) ranged from 92% [88%–94%] for 13q14 deletions with cell fractions below 5% to 71% [66%–76%] for cell fractions above 10% (**Fig. 3b**). 13q CN-LOH clones progressed to CLL at similar rates (**Fig. 3c**) despite being likely to have inactivated both copies of the 13q14 tumor suppressor region. This counterintuitive finding is consistent with epidemiological observations that CLL patients with monoallelic versus biallelic loss of 13q14 have similar prognosis^42,43^ and contrasts with effects of CN-LOH mutations that amplify myeloid CHIP mutations^44^.

The >2-fold larger number of UKB participants with a CLL-associated mCA (3,679 individuals with 13q14 deletion or trisomy 12) compared to the number with prevalent or incident CLL diagnoses (1,502 individuals) suggested that mCA calls might be a powerful tool for discovering germline variants that influence CLL risk. To evaluate the potential of this approach, we first verified that previously-identified CLL risk variants^45^ exhibited broadly consistent associations with CLL-associated mCAs (**Supplementary Fig. 5**). We then performed genome-wide association analysis on CLL-associated mCA status in UKB (i.e., presence of 13q14 deletion or trisomy 12) and on CLL case status in UKB. The mCA association analysis was better-powered, identifying 17 significant loci compared to 12, with 15 of the 17 loci falling within 500kb of a CLL risk locus found by a previous meta-analysis of 6,200 CLL cases^45^ (**Fig. 3d,e**). This result suggests that mCAs detectable from whole-genome sequencing of blood samples can provide more information about genetic predisposition to CLL than CLL outcome data from over a decade of follow-up.

Analyzing breakpoints of 13q14 deletions provided clues about the origins of these mutations and the varying strength of the clonal selection they experience. Discordant read pairs localized the breakpoints of 2,087 deletions in 1,590 individuals to ∼1kb resolution.

These breakpoints clustered in hotspots on the scale of 10–100kb (**Fig. 3f,g**), suggesting that local genomic or epigenomic architecture might strongly influence generation of mCAs. The most common left and right breakpoints of 13q14 deletions appear to be physically proximal in B-cells according to Hi-C contact maps^46^ (**Fig. 3f**), suggesting that chromatin looping in this region contributes to recurrent deletion of this 1Mb segment. Factors influencing clone fitness also appeared to shape the distribution of observed 13q14 deletion breakpoints. Shorter deletions tended to reach high cell fractions more often than longer deletions, suggesting that deletion of larger regions surrounding the locus may be detrimental to clone fitness (**Fig. 3g**). This effect, consistent with observations that mCA type is an incomplete determinant of clonal fitness^47^, remained after controlling for differential detection sensitivity for long versus short deletions by restricting to deletions with cell fractions >5% (**Fig. 3h**). Moreover, 13q14 deletions detected in conjunction with 13q CN-LOH tended to be shorter than 13q14 deletions without detectable CN-LOH (**Supplementary Fig. 4b**), hinting at selective pressures against biallelic deletions of larger regions that might contain essential genes^48^.

### CN-LOH mutations act on rare protein-altering variants in 38 genes

Copy-neutral loss of heterozygosity mutations do not alter gene dosage, yet are commonly observed in hematopoietic clones^3,6,7^ and comprised half of the mCAs we identified (**Fig. 1d**). We and others previously observed that some CN-LOH mutations appear to act on inherited or acquired protein-altering variants in several genes, conferring a proliferative advantage either by making a fitness-increasing variant homozygous or by removing a fitness-decreasing variant from a hematopoietic cell’s genome^21–23,32^. Here, new data from whole-genome sequencing of UKB—namely, our largest-to-date call set of 21,050 CN-LOH mutations, together with genotype calls for rare protein-altering SNVs and indels—provided the opportunity to find many more driver genes underlying CN-LOH expansions and study how these genes might influence the survival or proliferation of blood cells.

Protein-altering variants in 38 genes associated with strongly increased risk of CN-LOH mutations overlapping these genes (burden test *p*<1.2×10^-5^, FDR<0.01; **Fig. 4a** and **Supplementary Table 5**). Most of these associations were newly identified; for eight genes, inherited coding variants had previously been implicated as targets of CN-LOH mutations in UKB and BioBank Japan (BBJ)^21–23^, and for three genes (*DNMT3A*, *TET2*, and *JAK2*), we previously observed evidence of CN-LOH mutations acting on acquired CHIP driver mutations^22^. Gene ontology enrichment analysis of the 38 genes implicated DNA double strand break response (*p*=1.0×10^-7^), apoptosis (*p*=5.1×10^-6^), cytokine signaling (*p*=1.3×10^-6^), and protein ubiquitination (*p*=6.3×10^-4^) as cellular processes involved in clonal selection (**Fig. 4b** and **Supplementary Table 6**). Impaired DNA damage response can contribute to proliferation by enabling further accumulation of proliferation-increasing mutations or by removing replication checkpoints that ensure genomic integrity; cytokine signaling has important regulatory roles in immune cell proliferation; and ubiquitination regulates protein turnover, such that reduced function of ubiquitin ligases targeting pro-proliferative proteins could stimulate proliferation.

**Figure 4:**
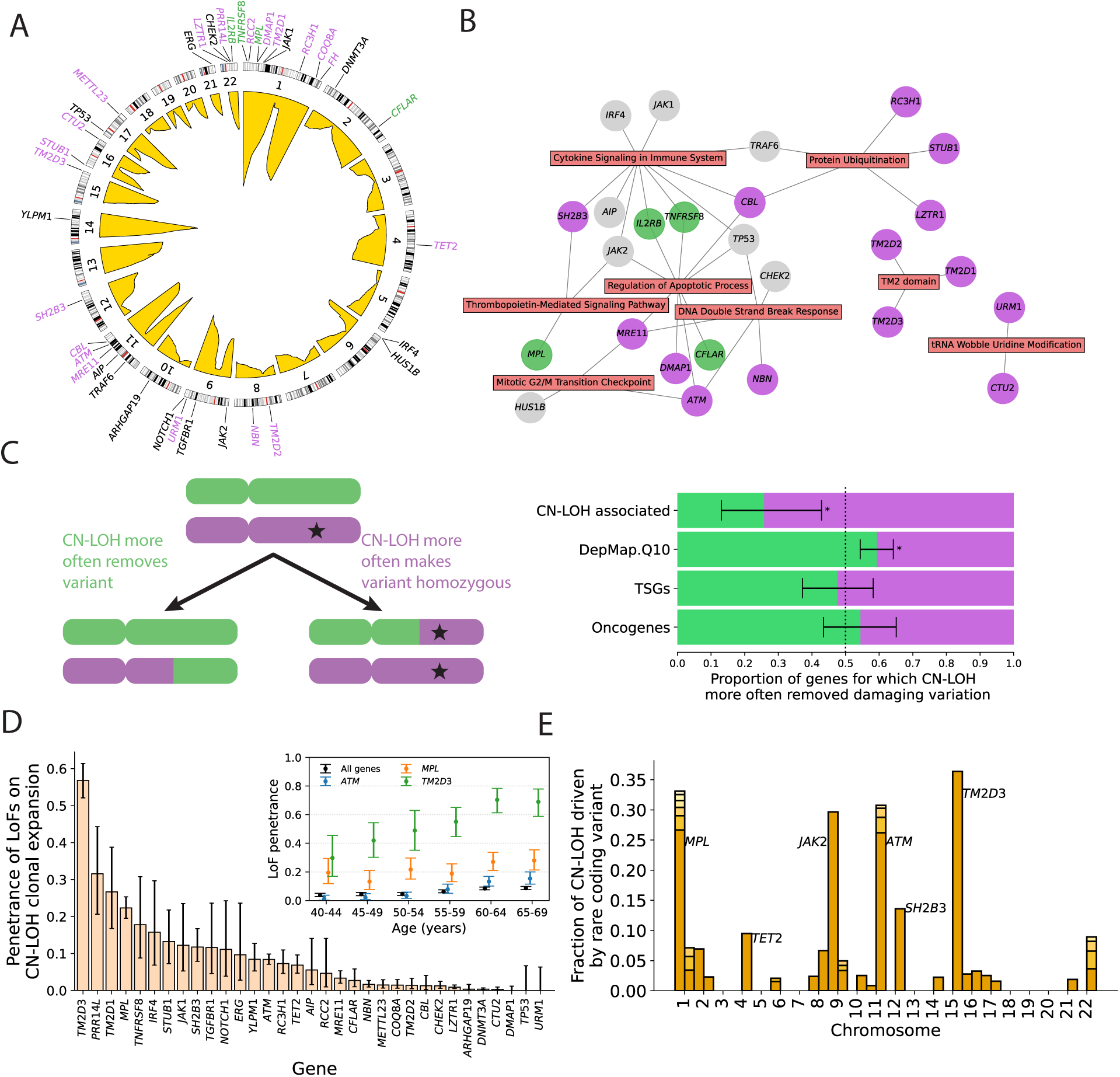
Rare protein-coding variants in 38 genes influence clonal expansion of CN-LOH mutations that modify their allelic dosage. **(A)** Genes for which a burden of rare coding variants associated with the presence of an overlapping CN-LOH mutation (*P*<1.2×10^-5^, FDR<0.01). Genomic coverage by CN-LOH mutations is shown in yellow (indicating relative frequencies of CN-LOH mutations on different chromosomes). Genes with coding variants that were more often made homozygous (resp., more often removed) by CN-LOH are colored purple (resp., green); genes for which a directional bias was unclear are colored black (**Methods**). **(B)** CN-LOH-associated genes cluster in DNA damage response, cytokine signaling, cell cycle regulation, and protein ubiquitination pathways. **(C)** CN-LOH mutations can either amplify or remove protein-altering variants. For each gene, we determined whether or not CN-LOH mutations observed in UKB more often removed LoF and high-impact (PrimateAI-3D score > 0.8) missense variants (based on a simple majority among carriers that could be phased). Proportions of genes for which coding variants were more often removed are shown in green for various categories of genes: DepMap.Q10, genes in the bottom decile of DepMap scores^59,60^; tumor suppressor genes (TSGs) and oncogenes are from MSK’s OncoKB^78^. Error bars, 95% Jeffreys intervals. **(D)** Penetrance of LoF variants (i.e., fraction of carriers for whom the corresponding mosaic CN-LOH mutation was observed) for 33 CN-LOH-associated genes with at least 30 carriers of rare (AF<0.001) LoF variants (in any transcript, and inclusive of LoF CNVs). Inset, penetrance as a function of age for *TM2D3*, *MPL*, *ATM*, and all 33 genes combined. **(E)** Fraction of individuals with CN-LOH detected on a given chromosome arm for whom the CN-LOH clonal expansion is potentially attributable to a rare coding variant (AF<0.01) in one of the 38 CN-LOH-associated genes (specifically, an LoF variant or a missense variant with PrimateAI-3D score >0.6 in any transcript).

The expanded list of putative targets of CN-LOH mutations also provided deeper insights into clonal selection on mutations in two previously-identified target genes, *TM2D3* and *CTU2*, which exhibit some of the strongest associations of protein-altering alleles with CN-LOH mutations in European and East Asian populations^21–23^. *TM2D3* encodes one of three transmembrane 2 (TM2) domain-containing proteins whose functions in human cells have yet to be characterized. Recent studies of model organisms have suggested that *TM2D* genes regulate Notch signaling^49,50^. Here, coding variants in *NOTCH1* and all three *TM2D* genes—*TM2D1*, *TM2D2*, and *TM2D3*—associated with CN-LOH mutations in *cis* (**Fig. 4a,b** and **Supplementary Table 5**), suggesting that these CN-LOH mutations may generate a proliferative advantage via dysregulation of *NOTCH1*, which is also a target of clonal expansion driver mutations (and CN-LOH) in esophageal tissues^51,52^. *CTU2* encodes a subunit of the cytosolic thiouridylase complex involved in maintenance of genomic integrity^53^. Here, we replicated the association of coding variation in *CTU2* with CN-LOH mutations in *cis* (previously identified in BBJ^23^) and found a new association of coding variants in *URM1* with CN-LOH mutations overlapping *URM1* (**Fig. 4b** and **Supplementary Table 5**). *URM1* encodes a ubiquitin-like protein that interacts with CTU2 in the thiolation of uridine in the wobble position of specific tRNAs^54^, suggesting a translation-related mechanism of clonal selection for these CN-LOH mutations.

For most CN-LOH mutations associated with a burden of protein-altering variants in a putative target gene in *cis*, haplotype phasing analyses enabled determining whether CN-LOH mutations more frequently made these variants homozygous or removed them from the genome (**Fig. 4c**). In our previous SNP-array-based analysis, CN-LOH mutations acted to make coding variants homozygous for all but one target gene, *MPL* being the sole exception^22^. Here, we again observed that for most putative target genes, CN-LOH mutations appeared to provide a “second hit”^55^ by increasing the dosage of a proliferation-increasing variant (**Fig. 4a-c**). However, we also observed four genes for which CN-LOH mutations generated revertant mosaicism, appearing to perform “natural gene therapy” on variants that presumably reduce cell fitness, replacing alleles carrying coding variants with unmutated alleles (**Fig. 4a**). These genes included *CFLAR*, which encodes a protein (CASPER, also called c-FLIP) that inhibits CD95-mediated apoptosis^56,57^, and *IL2RB*, which encodes a component of a receptor whose activation increases T cell proliferation^58^. This phenomenon appeared to extend more broadly to many other genes: among genes most essential for survival of lymphoid and myeloid cancer cell lines (lowest decile of DepMap gene dependency scores^59,60^), 225 genes experienced more CN-LOH mutations that removed damaging variants than CN-LOH mutations that made such variants homozygous, whereas only 154 genes had more CN-LOH mutations confer a second hit (two-sided binomial *p*=3.1×10^-4^; **Fig. 4c**). CN-LOH mutations also tended to more often duplicate LoF variants in tumor suppressor genes and more often remove LoF variants in oncogenes, as expected, though these differences did not reach statistical significance (**Fig. 4c**).

Loss-of-function (LoF) alleles of several putative target genes of CN-LOH mutations exhibited high penetrance for these CN-LOH clonal expansions (**Fig. 4d**). More than half of the UKB participants who carried *TM2D3* LoF variants had detectable 15q CN-LOH mutations overlapping *TM2D3*^21^, and *TM2D1* LoF variants exhibited the third-highest penetrance of 27% [17%–39%] (**Fig. 4d**). The penetrance of LoF variants associated with CN-LOH clonal expansions increased steadily with age (e.g., from 30% [17%–46%] to 69% [59%–78%] for carriers of *TM2D3* LoF variants of age 40–44 versus 65–69 years; **Fig. 4d**), reflecting the stochastic acquisition and slow clonal expansion of CN-LOH mutations over decades of aging^47,61^.

The 38 identified target genes of CN-LOH mutations provided plausible explanations for some CN-LOH mutations observed on 22 chromosome arms (**Fig. 4a,e**). However, while the newly-identified target genes provided insights into biological mechanisms that contribute to clonal hematopoiesis (**Fig. 4b**), they accounted for small fractions of the CN-LOH mutations observed on each arm—in contrast to the substantial fractions of mutations explained by target genes previously identifiable in smaller data sets^21,22^ (**Fig. 4e**)—suggesting that drivers other than rare coding variants in *cis* contribute to the expansions of most blood clones with CN-LOH mutations.

### Polygenic effects of common variants contribute modestly to CN-LOH expansions

Common genetic variation is another source of allelic differences between homologous chromosomes on which CN-LOH mutations can act to generate a proliferative advantage. Previous studies have identified four loci at which common variants associate with CN-LOH mutations in *cis*^22,23^. Here, after eliminating CN-LOH clonal expansions likely to be driven by rare coding variants, we observed just one previously-reported common-variant association at the *DLK1* locus^23^ (**Fig. 5a**). A genome-wide scan for common alleles associated with CN-LOH direction (i.e., a tendency for CN-LOH mutations in *cis* to make an allele homozygous, or alternatively, a tendency to remove it from the genome) identified four additional loci (*PRDM16*, *JAK2*, *ATM*, and *SH2B3*; **Supplementary Table 7**).

**Figure 5:**
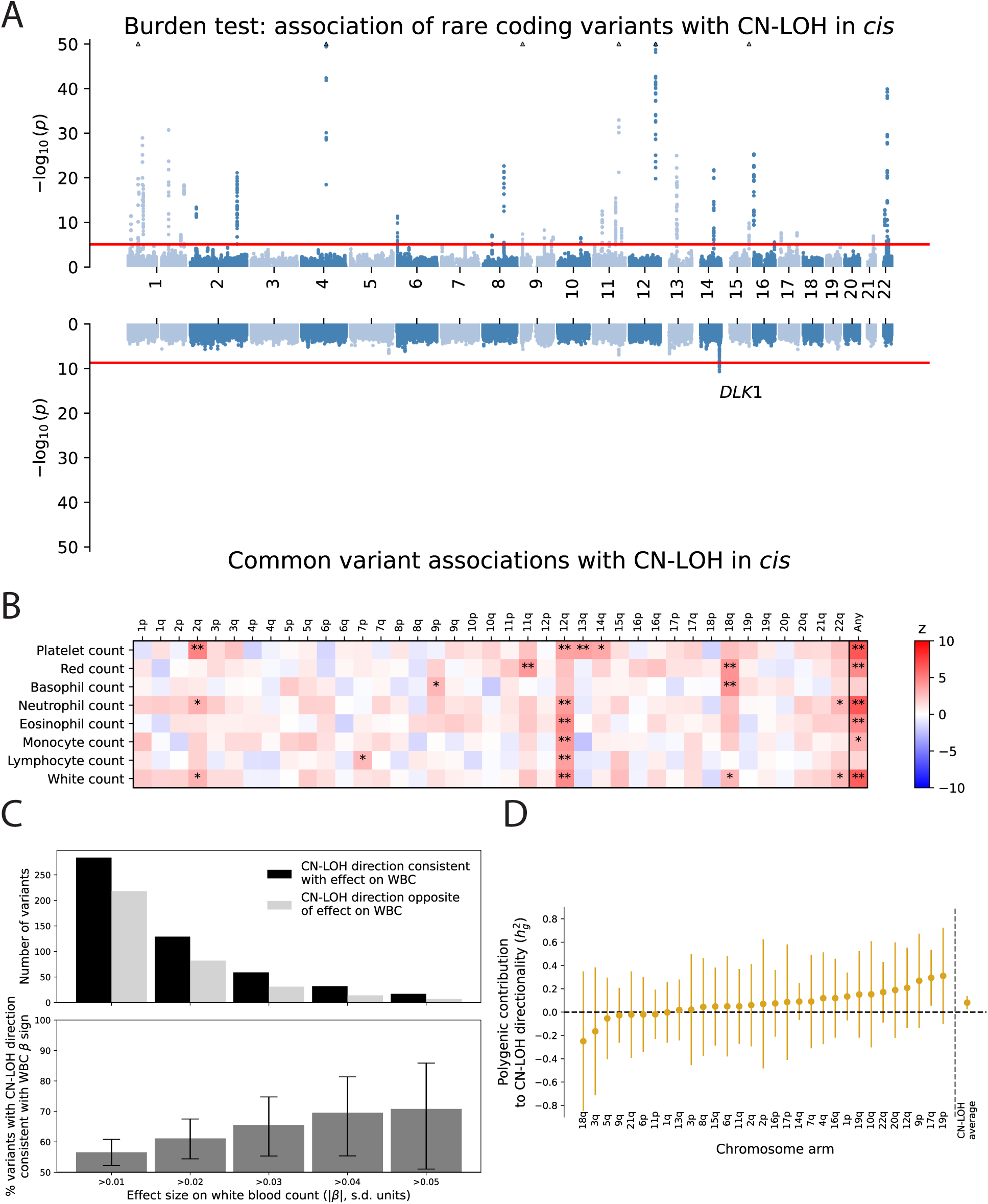
Polygenic contribution of inherited common variants to CN-LOH clonal expansions. **(A)** Miami plot comparing associations of rare coding variants (top; burden tests) and common variants (bottom) with presence of an overlapping CN-LOH mutation. *p*-values (y-axis; log_10_ scale) are from Fisher’s exact test. The significance threshold plotted for the burden test is the FDR<0.01 threshold of *p*<1.2×10^-5^, while the significance threshold for the common variant GWAS is the canonical threshold *p*<5×10^-8^. **(B)** Clonally expanded CN-LOH mutations in blood cells tend to amplify chromosome arms with proliferation-increasing polygenic scores for blood cell counts. The heatmap displays z-scores corresponding to the mean difference between polygenic scores on (i) haplotypes made homozygous versus (ii) haplotypes removed by CN-LOH mutations, divided by the s.e.m. Positive differences (red) correspond to chromosome arms for which the retained haplotype tended to have a larger blood count polygenic score than the removed haplotype. * FDR < 0.05; ** *P* < 0.05, Bonferroni-adjusted. **(C)** Alleles that associate with increased white blood count (WBC) tend to be more often made homozygous by CN-LOH. The effect directions of common WBC-associated index variants were evaluated for consistency with the directions of CN-LOH mutations that overlapped them (**Methods**). Error bars, 95% Jeffreys intervals. **(D)** Overall contribution of common inherited variants to determining the directionality of observed CN-LOH mutations (i.e., which haplotype is amplified and which is removed) estimated using variance components analysis (**Methods**). Estimates are shown for 31 chromosome arms with at least 200 CN-LOH mutations; the inverse-variance weighted average of these estimates is shown on the right. Error bars, 95% CIs.

Dysregulation of *PRDM16* has been implicated in myelodysplastic syndrome and acute myeloid leukemia involving t(1;3)(p36;q21) translocations^62^. The limited association signal from common variants contrasted strikingly with the many strong associations we observed between CN-LOH mutations and rare coding variants in *cis* (**Fig. 5a**).

Despite the paucity of common-variant associations with CN-LOH mutations, common variants could still influence *cis*-acting CN-LOH mutations at levels too subtle to discern at existing sample sizes (here, fewer than 1,000 CN-LOH mutations on most chromosome arms). We previously observed that differences in polygenic scores (PGS) for blood cell counts computed across alleles carried on haploid chromosome arms—proxying for differences in proliferative potential between pairs of homologous arms—correlated with CN-LOH directionality (i.e., which arm was duplicated and which arm was lost)^22^. This effect replicated and strengthened in significance here (**Fig. 5b**). Among specific variants associated with white blood counts (WBC), CN-LOH mutations in heterozygous carriers of a variant more often duplicated the WBC-increasing allele for most variants, and the strength of this effect increased with the magnitude of variants’ effect sizes on WBC (**Fig. 5c**).

Our CN-LOH call set also provided sufficient sample size to perform heritability analyses quantifying the extent to which the directions of CN-LOH mutations on a given chromosome arm are influenced by common variants spanned by these mutations. For these analyses, we defined a liability threshold model in which the observed direction of a CN-LOH mutation is given by the sign of an unobserved liability composed of a genetic component (namely, the difference in total proliferative strengths of the alleles carried on the two homologous chromosome arms) and a random non-inherited effect. We then used variance components analysis to estimate the fraction of variance in liability attributable to inherited common variants^63–65^. Averaging across chromosome arms with at least 200 CN-LOH mutations (excluding CN-LOH mutations containing rare coding variants in putative target genes), the average heritability attributable to common variants spanned by CN-LOH mutations was 0.08 (s.e.m. 0.03) (**Fig. 5d**). This modest common-variant heritability, together with the modest fraction of CN-LOH mutations (9%) that appear to act on rare coding variants in putative target genes, suggests that while allelic differences between the proliferative strengths of homologous chromosome arms play an important role in clonal expansions of many CN-LOH mutations, other proliferative influences must also drive expansion of clones with CN-LOH mutations. Further work will be required to determine whether CN-LOH mutations often represent passenger events on clones carrying other driver mutations, whether they act on epigenetic differences between homologous chromosomes, or whether they obtain proliferative advantages for other reasons.

## Discussion

In this study, we leveraged whole-genome sequencing of UK Biobank participants to obtain new insights into the landscape of mCAs in blood. Improved detection of short mosaic deletions revealed new hotspots driven by genome fragility and a surprisingly high prevalence of 13q14 mutations, while improved detection of large mCAs present at low cell fractions powered genetic association analyses identifying numerous targets of CN-LOH mutations.

These results show that because CN-LOH mutations commonly generate proliferative advantages by altering the dosages of inherited alleles, such mutations provide valuable clues that can point to fitness-influencing genetic variation. Consequently, increasingly large studies of CN-LOH mutations will provide increasing power to identify and fine-map further effects of inherited variants on clonal fitness, complementing analyses of somatic point mutations^66^. This contrasts with large mCAs that provide proliferative advantage primarily by altering gene copy number (e.g., trisomies), which provide non-specific information about their target genes no matter how many such mutations are observed.

The 38 genes that our burden tests identified as targets of CN-LOH mutations already provide intriguing insights into how these clones expand. For most of these genes, CN-LOH mutations provided a second hit, often to a gene in a proliferation-related pathway.

However, we also observed revertant mosaicism of damaging variants in several genes, and a broader analysis across DepMap-prioritized genes suggested that this phenomenon extends to dozens and perhaps hundreds of genes (based on the differential of 71 = 225 – 154 genes for which CN-LOH directionality favored reversion versus a second hit). This suggests that “natural gene therapy” to remove deleterious variants occurs much more frequently beyond previous observations in inherited bone marrow failure syndromes^67^, albeit to more modest extents in CH.

Our analyses of common-variant effects on CN-LOH mutations also demonstrate the promise of this approach: while we were not yet powered to identify many specific effects, we observed evidence of heritable, polygenic effects of common variants on CN-LOH directionality—suggesting the potential for future discovery from larger cohorts, as earlier analyses of complex trait heritability^65^ preceded a wave of GWAS discoveries.

Taken together, these results underscore that CN-LOH mutations have unique fitness consequences determined more by the alleles they carry than the chromosomes on which they fall^47^. Interpreting such mutations (e.g., to quantify risk of progression to malignancy) will require nuanced approaches. This need for nuance extends beyond CN-LOH mutations: even for 13q14 deletions, deletion length appeared to influence clonal fitness.

The surprising frequency of 13q14 deletion (>1% of 70-year-old individuals; highest among autosomal mCAs) allowed us to perform a natural history study of its progression to CLL. Given the prevalence of this mutation, an important area for future work will be to determine whether or how cases of mosaic del(13q14) should be clinically managed^68,69^: as with other forms of CH, the factors that determine which individuals progress to hematologic malignancy remain largely unknown. The high prevalence of 13q14 deletion also suggests the possibility that undetected 13q14 deletions and other mCAs could explain some cases of CH with unknown drivers, which is associated with risk of lymphoid disorders^15^.

Our analysis of UKB WGS data had several limitations: we could not study longitudinal dynamics^5^ of mCAs nor analyze specific cell populations of expanded clones^70,71^, nor could we study therapy-related CH^72,73^. Available sequencing depth also limited our ability to detect short mCAs with low cell fractions. Additionally, the UKB cohort had limited representation of non-European ancestries. Future WGS-based mCA analyses across multiple biobanks will provide opportunities to learn from other genetic ancestries, building upon recent SNP-array-based efforts^25,74,75^, and analyses of mCAs in other tissues^52,76,77^ will deepen our knowledge of the genetic forces shaping clonal selection more broadly.

## Acknowledgements

We thank V. Sankaran, S. Raychaudhuri, A. Gusev, A. Sekar, G. Genovese, and M. Hujoel for helpful discussions. This research was conducted using the UKB resource under application no. 40709. D.T. was supported by US NIH training grant T32 HG002295. N.K. was supported by US NIH training grant T32 HG002295 and fellowship F31 DE034283. R.E.M. was supported by US NIH grant K25 HL150334. S.R. was supported by a Swiss National Science Foundation Postdoc Mobility fellowship. P.-R.L. was supported by US NIH grants R56 HG012698, R01 HG013110, and UM1 DA058230 and a Burroughs Wellcome Fund Career Award at the Scientific Interfaces. The funders had no role in study design, data collection and analysis, decision to publish or preparation of the manuscript. Computational analyses were performed on the O2 High Performance Compute Cluster, supported by the Research Computing Group, at Harvard Medical School (http://rc.hms.harvard.edu) and the UK Biobank Research Analysis Platform.

## Author Contributions

D.T. and P.-R.L. performed analyses and wrote the manuscript. N.K., R.E.M., and S.R. provided guidance on the analyses and interpretations of results.

## Data Availability

Full summary association statistics for the four GWAS in Supplementary Fig. 6 will be released in a Zenodo repository prior to publication. WGS read-depth principal components and empirically estimated REF-bias in allelic depths at common SNPs and indels are provided at https://data.broadinstitute.org/lohlab/mCAs_WGS/. Individual-level mCA calls will be returned to UK Biobank for release on the UKB-RAP platform. Access to UK Biobank (http://www.ukbiobank.ac.uk/) is obtained by application.

## Code Availability

Custom code used to call mCAs and perform downstream analyses can be found at https://github.com/tangdavid/mCAs_WGS. The following open-source software packages were also used: samtools v1.15.1, bcftools v1.15.1, plink v1.9, plink v2.0, IMPUTE5 v2.0.0, XCFtools v5.0.0, SHAPEIT5 v5.1.1, GCTA v1.94, REGENIE v3.1.1, python 3.8.10.

## Methods

### Ethics

This research complies with all relevant ethical regulations. The study protocol was determined to be not human subjects research by the Broad Institute Office of Research Subject Protection and the Partners HealthCare Human Research Committee (as all data analyzed were previously collected and de-identified).

### UK Biobank data set

UK Biobank is a prospective cohort of half a million volunteer participants of age 40–70 years at recruitment between 2006–2010. Blood samples were acquired at initial assessment from which aliquots were used for SNP-array genotyping, serum biochemistry assays, and blood count assays^35^, with remaining blood preserved for future analyses.

Health-related phenotypes were recorded from touchscreen questionnaires and nurse interviews, and follow-up health outcome data has been accruing from linkage with UK national health registries.

WGS data was subsequently generated and made available for 490,414 UKB participants sequenced by the UK Biobank Whole Genome Sequencing Consortium^36^. Blood-derived DNA was sequenced using Illumina NovaSeq 6000 machines to an average coverage of 32.5x, after which 151bp paired-end reads were aligned to GRCh38 and SNP and indel calling was performed as previously described^36^. Blood samples used for WGS had been acquired at initial assessment for 99.6% of sequenced individuals (and for 99.7% of the 484,081 individuals in our primary data set); for the remaining individuals, blood samples from a later visit to a UK Biobank assessment center were used for sequencing. Given the small fraction of participants whose sequenced blood samples had been obtained after initial assessment, we included all individuals in our analyses (using age at initial assessment in analyses of age; averaged across the cohort, this underestimated age at blood draw by ∼1 week).

UK Biobank participants can withdraw from the study at any time and request that their data no longer be used. Consequently, some analyses reported in this manuscript have small discrepancies in participant counts, reflecting slight variability in the number of available (non-withdrawn) participants at the time of each analysis.

### Overview of approach for detecting mCAs from WGS data

An effective approach for detecting mCAs from bulk genotyping or sequencing data is to search for long (megabase-scale) stretches of genome in which the allelic fractions of an individual’s two haplotypes exhibit imbalance^21,79^. This approach can be applied to either SNP-array genotyping intensity data or WGS data by appropriately modeling allelic imbalance information derived from either of these sources. Most large-scale studies of mCAs performed in the past several years have analyzed SNP-array data using the MoChA software package^21,22^ to model B-allele frequency (BAF) measurements computed from signal intensities of allele-specific genotyping probes. Recently, MoChA has also been used to perform WGS-based mCA calling by analyzing imbalances in “allelic depths” computed from counts of haplotype-informative sequencing reads that span a heterozygous site^29^.

Compared to SNP-array data, high-coverage (∼30x) WGS data can in theory provide more information about allelic imbalance—and enable higher-resolution mCA calls—because WGS data contains information about allelic balance at most heterozygous sites in a human genome (typically 2–3 million) rather than only those represented on a SNP-array (typically 100,000–200,000 hets per genome). This difference in the number of informative measurements is only partially offset by SNP-array intensity data tending to provide more information per heterozygous site (i.e., lower s.d.(BAF), typically 0.03–0.07) than WGS allelic depths.

A recent analysis of the TOPMed WGS data set using MoChA demonstrated promising results of WGS-based mCA detection^29^: autosomal mCAs were detected at rates similar to the rates we previously observed in analyses of UKB SNP-array data (for individuals in corresponding age ranges)^22^. This similar detection sensitivity suggests that the higher information content of WGS compensated for the lower phasing accuracy expected in the smaller, more diverse TOPMed cohort.

Here, we sought to further enhance mCA detection in the UKB 500K WGS cohort by performing an optimized analysis of this very large WGS data set that addressed a few key limitations of MoChA’s WGS-based mCA analysis pipeline. First, we performed genome-wide copy-number profiling on each WGS sample to (a) flag and filter individual-specific genomic regions potentially containing inherited copy-number variants and (b) accurately calibrate WGS read-depth measurements. Germline structural variants (particularly inherited duplications) are a major source of false positives in mCA analysis, such that stringent post-processing of mCA calls from existing pipelines is needed to filter false-positive mosaic calls^29^. Additionally, technical noise in WGS read-depth measurement limits the extent to which copy-number states of mCAs can be confidently determined.

Second, we recomputed allelic depths at heterozygous sites in a way that avoids double-counting allelic observations derived from the same DNA fragment. Because paired-end sequencing reads can overlap multiple nearby heterozygous sites, standard allelic depth measurements (which count the total number of sequencing reads supporting each allele) at nearby heterozygous sites do not provide independent measurements of allelic imbalance (**Supplementary Fig. 1**). MoChA circumvents this issue by imposing a minimum genomic distance between heterozygous sites considered in allelic imbalance analysis, dropping allelic depth measurements from sites that are deemed to be too near other sites. This approach loses some information, so here we instead “de-duped” allelic depths by examining the read pair from which each observed allele is derived to generate allelic depths derived from nonredundant observations that we could subsequently model as independent. These analyses are described in detail below.

### Genome-wide copy-number profiling and WGS read-depth calibration

WGS depth-of-coverage (i.e., read-depth) analysis has long been used to detect genomic copy-number variants^80–82^. CNVs manifest as local increases or decreases in a sample’s genomic read-depth profile, as the number of reads observed to align to a given genomic region scales with copy number of the region. However, a challenge of read-depth-based CNV analysis is that technical biases in whole-genome sequencing can also produce local variation in read-depth (unrelated to copy-number variation) that can exhibit different patterns in different samples. CNV analysis methods have typically accounted for these biases by modeling how a sample’s read-depth varies as a function of local sequence properties (e.g., GC content) and/or comparing a sample’s read-depth profile to that of other samples (ideally samples sequenced in the same batch). These approaches for normalizing WGS read-depth measurements have been effective for calling germline CNVs, which increase or decrease copy number by 50% or more (CN=2 to CN≤1 or CN≥3).

However, for the purpose of detecting mCAs and classifying their copy-number states (deletion, CN-LOH, or duplication), we needed to develop a copy-number profiling pipeline that satisfied a different set of considerations. Because mCAs in healthy blood often have low clonal fractions (on the order of ∼1% or lower) but can span entire chromosomes, we needed an approach that could (i) conservatively identify and filter any genomic regions that might contain germline CNVs (because such CNVs, even if relatively small, could contribute noticeable noise in read-depth relative to the read-depth changes produced by large but low-cell-fraction duplications or deletions); and (ii) normalize WGS read-depth measurements as precisely as possible, ideally reducing the effects of residual technical biases to well below 1%. To achieve these goals, we developed a new copy-number profiling approach that involved two main steps, detailed below: (1) GC-bias correction and CNV-masking, and (2) principal component analyses (PCA)-based denoising.

#### GC-bias correction and CNV-masking

For each WGS sample, we first computed an empirical GC profile following the approach of Genome STRiP v2^83^ with a few optimizations to improve computational efficiency. Briefly, for each possible value of local GC content (discretized to GC% = 0%, 1%, 2%, …, 100%), we measured the average number of read pairs aligned at genomic positions with that GC% in a surrounding 400bp window. We considered a read pair to align at the alignment start position in GRCh38 if the read mapped on the right. We excluded genomic positions in regions with low complexity, low mappability, and common CNVs (specifically, the union of Genome STRiP’s lcmask, svmask, and gcmask for hg38, together with common CNVs (global AF>1% or EUR AF>1%) in the 1000 Genomes 30x SV call set^84^ or gnomAD-SV v2.1^85^). To perform this analysis efficiently, we pre-generated a binary file containing genome-wide GC content (in sliding 400bp windows) and genome mask status across GRCh38, downsampled to 32bp resolution. For each sample (i.e., WGS cram file), we then performed a single-pass analysis in which we tabulated GC profile statistics and simultaneously recorded the number of read pairs aligned to each 1kb bin of GRCh38 (applying the same set of genomic masks except the Genome STRiP gcmask). We stored this information in a compact 1kb-resolution read-count profile (counting one read per pair) for downstream processing. This computation required ∼8 minutes per 30x WGS sample on a single vCPU, making analysis across the full UKB cohort feasible.

To identify potential germline CNVs (to mask from downstream analyses), we then used a simple three-state HMM (deletion, no CNV, duplication) to identify genomic segments (i.e., sequences of 1kb bins) with read counts that indicated potential copy-number variation.

Specifically, for a given sample, we computed the expected read count within each 1kb bin based on local GC content (in sliding 400bp windows overlapping the bin) and the GC profile of that sample, excluding masked regions. For each chromosome, we computed an adjustment to this expected read count based on the median ratio (across 1kb bins on the chromosome) of observed to expected reads. For each 1kb bin, for each HMM state, we then computed the HMM emission probability for that copy-number state using a Gaussian approximation to a Poisson model (i.e., mean and variance both equal to the expected read count accounting for the assumed copy-number state). We set the HMM jump probabilities to 0.001 for transitions between unequal states, and we called potential CNVs using the Viterbi algorithm.

Compared to typical germline CNV callers, this simple HMM was optimized for high sensitivity because our primary goal was to exclude any regions potentially containing kilobase-or-larger CNVs from mCA analysis. As such, false-positive CNV calls were acceptable given that they constituted a conservative error (in the context of mCA-calling) that would only slightly diminish the set of genomic regions available for analysis of a given sample. The MoChA pipeline also includes a CNV-masking step but does so using allelic depth (AD) information at heterozygous sites; here, considering all observed reads (including those that do not overlap heterozygous sites) increases sensitivity to detect and filter germline CNVs.

#### Denoising WGS read-depth profiles using PCA

While GC profile correction is reasonably effective at adjusting WGS read-depth measurements for effects of sequencing biases that correlate (at least in part) with local GC content of genomic regions, we observed that even after GC correction, normalized read-depth profiles (observed / expected) exhibited residual “waves” at megabase and sub-megabase scales with amplitudes on the order of 1%. These genomic waves varied from sample to sample, such that we could not simply compute and correct for an average wave pattern observed across samples. However, we reasoned that these wave patterns might nonetheless be driven by a relatively small set of technical and biological factors (e.g., sequencing chemistry, library preparation, relative representation of blood cell types, DNA quality, etc.). If such factors each affected different samples to different extents, then we could potentially use principal component analysis to estimate the key components of read-depth variation, after which we could correct for the estimated contributions of these components to a given sample’s genome-wide read-depth profile. Such an approach could effectively learn and adjust for effects of unknown covariates (by observing and then decomposing their combined effects across many samples) without having to specifically identify these covariates.

To implement this approach, we first needed to compute the top PCs of a set of normalized read-depth profiles observed across a large reference set of WGS samples that were minimally affected by mCAs (as copy-altering mCAs present in many samples could infiltrate PCs). We first computed genome-wide read-depth profiles for all WGS samples at ∼20kb resolution (reasoning that this resolution was coarse enough for read-depth measurements to have sufficient signal-to-noise to be informative of wave patterns, yet fine enough to adequately model genomic waves). To do so, we first partitioned GRCh38 into chunks containing ∼20kb of base pairs that were not masked from read-depth analysis (such that masked regions expanded the physical span of chunks). For each WGS sample, for each chunk, we then computed the ratio of the observed to the expected number of reads aligned to the chunk, using the sample’s GC profile to compute the expected number of reads, and masking any portion of the chunk that overlapped germline CNV calls from the HMM (described above). This vector of per-chunk observed / expected ratios constituted the sample’s normalized read-depth profile.

We then selected samples for inclusion in the reference set (for PC computation) based on the following initial filters: age at most 41 years old at recruitment, no SNP-array-based mCA call^22^ (on any autosome or sex chromosome), and sample contamination at most 0.002 (as estimated by DRAGEN). We further required that samples have read-depth-based ploidy close to expectation (median(normalized read-depth) within 1±0.03 for autosomes, within 1±0.02 for chrX in females, and within 0.5±0.02 for both chrX and chrY in males). Finally, we excluded any female samples with male contamination >0.0005 based on chrY normalized read-depth.

We split the reference cohort by sex and computed normalized read-depth PCs separately for the male (XY) and female (XX) subsets. For each sex, we first computed a “baseline” profile across samples (which was very close to 1 for most genomic chunks). We computed this baseline to have the property that dividing each sample’s normalized read-depth profile vector by this baseline vector (element-wise) and cropping values to 1±0.08 (to reduce the influence of outliers) produced an average value of 1 for each chunk (averaging across reference samples). We then obtained PCs from the singular vector decomposition (SVD) of the matrix comprised of the resulting baseline-normalized, cropped read-depth profile vectors (after subtracting 1 to recenter values to 0). Finally, we applied one additional step to limit the extent to which mosaic sex chromosome loss could infiltrate these PCs: we took the top *k* PCs for which these PCs cumulatively explained no more than 5% of variance of a sex-chromosome-loss indicator vector (X loss for females, Y loss for males), after which we required that subsequent PCs be orthogonal to the sex-chromosome-loss vector. To achieve this orthogonality, we projected out the top *k* PCs and the sex-chromosome-loss indicator vector from the read-depth matrix and computed the SVD of this matrix to obtain top subsequent PCs.

The above procedure generated, for each sex, a baseline vector and a set of top PCs. To use this information to compute PC-adjusted normalized read-depths for a given WGS sample (including samples not in the reference set), we first computed the sample’s baseline-normalized, cropped read-depth profile (in the same way as above) and then computed its components along the top 20 PCs (obtaining the PC coefficients by computing inner products with the sample’s profile). This generated a sample-specific PC-derived genomic wave correction that we could then use to adjust the expected read count in each 1kb bin of GRCh38:

Finally, we applied one additional adjustment to handle potential aneuploidies (from very large inherited or mosaic CNVs): we computed the median per-chromosome depth across autosomes and computed a final calibration factor assuming that this value should correspond to copy number 2.

In this way, we obtained sample-specific GC-corrected, PC-corrected, aneuploidy-corrected values of expected read counts across 1kb bins of the genome that we could then use to compute normalized read-depth in mCA regions (by taking the ratio of observed to expected reads).

### Generating phased haplotypes of common variants in UKB

To effectively use WGS data to assess allelic imbalance across the heterozygous SNPs and indels throughout each UKB participant’s genome, we needed to generate phased haplotypes that would allow assigning each sequencing read spanning a heterozygous variant to one haplotype or the other. To do so efficiently and accurately, we started with the SNP-array scaffold of phased haplotypes that we had previously generated for UKB using Eagle2^22,86^ and used imputation to extend it to common SNPs and indels (MAF>0.01) not in the scaffold. Specifically, we first lifted the SNP-array scaffold to GRCh38, dropping variants that lifted to out-of-order positions or to positions with different reference bases in GRCh38 vs. GRCh37. We then used IMPUTE5^87^ to impute 10.0 million autosomal common variants (MAF > 0.01) from the UKB 200K WGS SHAPEIT5-phased haplotype reference panel^88^ onto the SNP-array scaffold haplotypes (available for 484,149 participants with WGS data after excluding participants who had withdrawn). Because common variants could be accurately imputed without using the full sample size of the 200K WGS reference panel, we split the panel into two equal halves and performed imputation using only half the panel (approximately 100,000 individuals). This approach also allowed us to perform imputation in an out-of-sample manner: for individuals represented in the panel, we used each half of the reference panel to impute into our SNP-array scaffold haplotypes for individuals in the other half of the panel. This out-of-sample imputation enabled a QC check in which we masked individual-level variant calls for which the imputed genotype disagreed with the (unphased) WGS-derived genotype (from the DRAGEN single-sample variant call files released by UKB), leaving heterozygous SNP and indel calls with one REF and one ALT allele that were consistently called from WGS and imputed from the reference panel.

To minimize the presence of heterozygous variant calls with allelic balances affected by inherited structural variation, we also masked variants that fell in potential germline duplications identified from the genome-wide copy-number profiling procedure described above. Across all individuals, we additionally excluded all variants in genomic blacklists consisting of low complexity regions, common structural variants, and multiallelic copy number variants (as described above). This pipeline produced high-quality phased haplotypes of 9.3 million common variants for 484,149 UKB participants with both WGS data and SNP-array scaffold haplotypes.

### Recomputing allelic depths and estimating REF-bias of common SNPs and indels

The DRAGEN single-sample VCF files provided by UKB contained allelic depth (AD) information that was not ideal for allelic imbalance analysis because of the issue of ADs at nearby heterozygous sites double-counting information derived from the same DNA fragment. To generate “de-duped” allelic depths that only counted a given DNA fragment (i.e., read pair) in at most one AD measurement, we ran *samtools mpileup*^89^ on individual-level sequencing read alignment (CRAM) files to extract the read names of sequencing reads spanning heterozygous sites. We then recomputed ADs using only sequencing read names that had not already contributed to an AD at a previous heterozygous site. Because paired-end sequencing reads share the same read name, this procedure ensured that each DNA fragment was only counted once.

An additional challenge of modeling allelic imbalance based on WGS-derived allelic depths is that read alignments at a heterozygous site tend to exhibit a bias in favor of the reference allele (REF) because non-reference alleles incur mismatch penalties in read-mapping algorithms, reducing the rate at which read alignments supporting non-REF alleles are observed. To enable modeling of this REF-bias during mCA-calling, we estimated the mean REF-bias of each common variant in the data set based on de-duped ADs across 10,779 UKB participants (restricting the REF-bias computation to the subset of individuals heterozygous for each variant). We filtered 1.7% of common SNPs and 62.6% of common indels for which the REF-bias exceeded a threshold of 0.05, reasoning that ADs for these variants might exhibit too much bias to allow robust allelic imbalance analysis.

### Calling mCAs and assigning copy-number states

To call mCAs using de-duped allelic depths and the phasing information described above, we reimplemented the BAF+phase hidden Markov model (HMM) from MoChA, which models phased allelic fractions at consecutive heterozygous sites (assuming a model in which chromosomal segments can exhibit varying amounts of allelic imbalance 𝜃, where 𝜃 is a hidden state parameter)^21,22^. Specifically, we modified the emission probabilities of the HMM (which model observed allelic depths of the REF and ALT alleles at heterozygous variants) to account for the level of REF-bias we estimated for each variant. If 𝑎 quantifies the allelic fraction of haplotype 1 in the sampled cell population (such that 𝑎=0.5±𝜃, and 𝑎≠0.5 corresponds to the presence of an mCA) and 𝑏 denotes the fraction of mapped reads that support the REF allele in non-mosaic heterozygous individuals (i.e., the estimated amount of REF-bias; typically 𝑏>0.5), then assuming 𝑛 reads are observed in total (i.e., 𝑛 is the sum of the REF and ALT allelic depths), the number of reads supporting the REF allele, 𝑋, follows the binomial distribution 𝑋 ∼ 𝐵𝑖𝑛𝑜𝑚𝑖𝑎𝑙(𝑝, 𝑛), where 𝑝 is given by the following formula (depending on whether or not the REF allele is on haplotype 1 or haplotype 2):

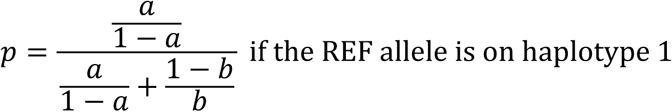

and

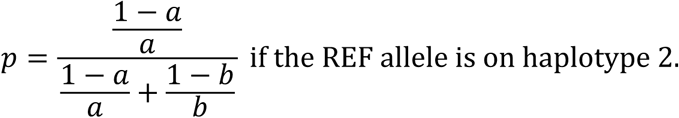

Additionally, to optimize the HMM for calling mCAs in a large WGS cohort, we adjusted the HMM transition probabilities and the set of hidden states considered by the HMM. Specifically, we set the mCA start and stop transition probabilities to 10^-5^ and 10^-4^, respectively, to reflect increased detection sensitivity in WGS, and we set the phase switch probability to 10^-4^ to reflect the accurate long range statistical phasing in UKB^22^. We also increased the representation of both low and high levels of allelic imbalance in the discretized set of hidden states 𝜃 considered by the HMM, discretizing 1/𝜃 into the following list: 4, 5, 6, 8, 10, 15, 20, 30, 50, 80, 120, 145, 170, 230, 310, 400, 500, 610. (The precise choice of discretization is less important than the range of imbalance amounts considered, because after an mCA is called at a sequence nonzero states 𝜃 in the Viterbi path through the HMM, the allelic imbalance within the mCA is recomputed without discretization.) We chose these parameters to maximize detection rate while maintaining high levels of WES validation in a pilot analysis of 10,774 randomly selected individuals (see **Validation of mCA calls using whole-exome sequencing data** below).

Finally, for each mCA call, we computed the relative WGS read-depth within its boundaries (i.e., the fractional amount of increase or decrease in read-depth relative to expectation assuming diploid copy number) and provisionally classified each mCA as a loss if the relative depth was less than -0.6×(allelic imbalance), a gain if the relative depth was more than 0.8×(allelic imbalance), and a CN-LOH otherwise (Fig. 1c and **Supplementary Fig. 2a**). Due to difficulties calibrating read-depth in genomic regions that undergo somatic V(D)J recombination during B and T cell maturation (such that mosaic V(D)J deletions are expected to cause leukocyte-derived WGS data to have average copy number less than 2 in these regions), we reclassified all interstitial mCAs that had been classified as CN-LOH with boundaries between chr2:85-92Mb, chr7:35-40Mb, chr7:140-145Mb, chr14:20-25Mb, and chr22:20-25Mb as loss, reasoning that these mCA calls most likely reflected clonal expansions of cells that had experienced deletions in the BCR or TCR regions during V(D)J recombination. We also reclassified a subset of erroneous full chromosome CN-LOH calls as isochromosomes (which involve the simultaneous loss of one chromosome arm and gain of the other) if the product of the depth z-scores on the p-and q-arms for a given call was less than -9.

### Prefiltering to optimize computational efficiency

Because retrieving the read names of sequencing reads from CRAM files is computationally demanding to perform on hundreds of thousands of WGS samples, we implemented a prefiltering strategy to efficiently nominate a set of chromosomes (on average 1 autosome per individual) that potentially harbored an mCA. We then performed full analysis only on this smaller set of nominated chromosomes.

For the prefiltering step, we ran MoChA with permissive parameters on the allelic depths (ADs) provided in the DRAGEN single-sample VCF files: specifically, we limited masking of heterozygous sites close to other heterozygous sites (by setting *--min-dist* to 50), disabled the LRR model (with *--LRR-GC-order* 0, *--LRR-weight* 0, *--bdev-LRR-BAF* 6),and set custom transition probabilities in the HMM (*--auto-tel-pl* 20, *--xy-major-pl* 50, *--xy-minor-pl* 40, *--ffip-pl* 40, corresponding to the choices we used in the full analysis). This prefiltering run of MoChA was designed to have low specificity (as it ignored double-counting of fragments overlapping two or more heterozygous sites) but high sensitivity, prioritizing an average of 4.4% of the data set—i.e., any chromosome on which MoChA found potential evidence of an mCA—for thorough analysis.

This prefiltering pipeline had a minimal impact on mCA detection sensitivity: in a pilot analysis of 10,779 participants for whom we performed full analysis on all chromosomes, the set of chromosomes nominated by prefiltering included 98.5% of the mCA calls identified by full analysis.

### Post-processing mCA calls

Because our genome-wide copy-number profiling procedure had largely eliminated confounding allelic imbalance signal from inherited copy-number variation, our mCA call set required minimal post-processing. We first dropped eight very short (<2kb) calls for which we were unable to obtain accurate read-depth estimates. We then filtered out a total of 256 short gains (<1Mb) called at chr4:126Mb, chr4:168Mb, chr7:101Mb, chr8:4Mb, chr8:86Mb, chr14:22Mb, and chr14:35Mb that appeared to arise from uncaught germline duplications based on manual inspection of their breakpoints and their estimated cell fractions (as germline duplications manifest as clusters of “gains” that are all at near-100% clonal fraction and all share similar breakpoints). We also filtered 38 mCA calls at chr4:39-40Mb and 27 mCA calls at chr10:111-112Mb (predominantly loss calls at both loci) that appeared to be technical artifacts arising from large repeat expansions of at *RFC1* and *FRA10B*, respectively; patterns of read-depth deviation at these loci suggested that large repeat expansions might have caused local distortions of allelic depths in the regions flanking these repeats. We also filtered 25 mCAs called on chromosome 21 in 10 individuals with constitutional trisomy 21. Finally, we applied two sample-level filters to remove 64 individuals with DRAGEN-estimated sample contamination >0.02 (among whom 30 mCAs had been called) and four individuals with >50 interstitial CN-LOH calls (among whom 337 mCAs had been called). These filters produced our final call set of 43,617 autosomal mCAs called in 484,081 UKB participants.

### Validation of mCA calls using whole-exome sequencing data

The high-level characteristics of our mCA call set—the genomic distribution of mCAs (Fig. 1b), relationship of read-depth to allelic imbalance (Fig. 1c), and enrichment of mCAs with age (Fig. 1d)—provided strong evidence that our analytical pipeline had produced robust mCA calls. To obtain additional confirmatory evidence, we analyzed whole-exome sequencing (WES) data previously generated for most UKB participants^37^ to determine whether allelic imbalances within mCAs called from WGS data replicated in WES data. We extracted de-duped allelic depths from the WES cram files with the same samtools mpileup pipeline that we used on the WGS cram files and then downsampled the allelic depths to adjust for WES REF-bias, which we empirically estimated from a subset of 10,779 WES cram files. For each mCA called from WGS for which WES data was available, we computed the total allelic depth (across heterozygous sites within the mCA) supporting each of an individual’s two haplotypes, accounting for any phase switch errors inferred by the HMM. We then tested whether the direction of allelic imbalance in the WES ADs agreed with the direction determined from WGS. Because many mCAs had limited coverage by WES reads, we expected the direction to sometimes disagree by chance. We quantified the expected probability of directional concordance per mCA and observed good agreement between expected and observed rates of concordance (**Supplementary Fig. 2b**). Specifically, to quantify the expected probability of directional concordance, we modeled the number of WES reads from the overrepresented haplotype as normally distributed with mean 𝑛(0.5 + 𝜃̂) and variance 𝑛(0.5 + 𝜃̂)W0.5 − 𝜃̂X + 𝑛(𝑛 − 1)𝜎_𝜃_^2^ where 𝜃̂ is the estimated allelic imbalance within the mCA, 𝜎̂_𝜃_ is the standard error of the estimated allelic imbalance (such that the second term in the variance formula accounts for uncertainty in this WGS-based estimate), and 𝑛 is the total number of WES reads that mapped within the called mCA. We took the complement of the CDF evaluated at 𝑛/2 as the expected concordance.

### Comparison to MoChA

To evaluate the extent to which our optimized analysis of the UKB 500K WGS data set improved mCA detection performance, we compared mCA calls from the pipeline to calls obtained using MoChA in a pilot analysis of 10,744 individuals. First, we ran MoChA with default parameters, providing as input ADs from the DRAGEN VCF files and the same phased haplotypes that we used in our main analysis. This run generated a raw output file containing an average of 2.97 autosomal mCA calls per individual; this extremely large call rate indicated that stringent post-processing was needed to filter false positives. Applying the post-processing filters suggested in the MoChA documentation reduced the call set to 551 autosomal mCA calls (0.05 per individual). In comparison, our pipeline generated 942 autosomal mCA calls for these individuals, of which approximately 422 were shared (based on affecting the same chromosome in the same individual). The mCA calls unique to the MoChA analysis appeared likely to have a high false positive rate given weaker age skew in these individuals (mean age 58.0 (s.e.m. 0.7) compared to mean age 59.1 (s.e.m. 0.4) for individuals with mCAs called by both pipelines; the mean age at assessment across all UKB participants was 56.5 years). Among mCA calls shared between the MoChA analysis and our analysis, the amount of allelic imbalance information supporting an mCA (quantified by LOD score; log_10_ odds) tended to be ∼1.2-fold higher in our analysis, consistent with our read-level analyses recovering information that had been lost by MoChA’s thinning of heterozygous variants to a minimum separation of 400bp.

### Defining focal mCA regions

We defined focal regions by scanning the genome in 1Mb windows and counting (separately) the numbers of short (<2Mb) mosaic deletions and duplications that either partially or fully overlapped each 1Mb window. We considered windows with more than 10 mCAs to be focal regions and merged adjacent windows to obtain our final list of 53 regions. For each region, we calculated a focal index as the maximum number of individuals with an mCA overlapping a 1kb bin within the focal region (maximizing across all possible choices of this 1kb bin) divided by the total number of individuals with an mCA overlapping any part of the focal region. We annotated focal regions as fragile sites in cancer based on the 19 autosomal fragile loci described in the somatic structural variant analysis of the Pan-Cancer Analysis of Whole Genomes (PCAWG) Consortium^38^.

### Localizing mCA breakpoints using discordant reads

Because allelic imbalance information derived from WGS read counts is noisy, mCA breakpoints can only be approximately estimated from allelic depth data, such that true breakpoints may be tens of kilobases away even for mCAs with high clonal fractions. We therefore analyzed discordantly-mapped read pairs (which can arise when a sequenced DNA fragment from a cell containing the mosaic mutation spans an mCA breakpoint) to better localize the breakpoints of a subset of mCAs. For each individual with at least 1 and fewer than 20 mCA calls, we identified WGS read pairs for which both reads mapped within 50kb of a called mCA breakpoint, filtering out read pairs that both mapped to the same chromosome with an insert size less than 5kb. We binned the remaining “discordant” read pairs by truncating the genomic position of both the read and its mate to the nearest kb, and we recorded the number of read pairs observed in each bin-pair as well as orientation (i.e., strand) information for the read pairs.

To find discordant read pairs originating from simple deletions and tandem duplications, we restricted to discordant read pairs mapping to the same chromosome. Among bin-pairs containing such discordant reads, we merged adjacent bin-pairs (for which the left-side bins were either the same or differed by 1kb, and likewise for the right-side bins) and then restricted to bin-pairs for which all discordant read pairs had the same orientation consistent with either a deletion or a duplication. For deletions, the left read in a pair should map in the forward direction and the right read in a pair should map in the reverse direction, and for duplications, the read orientations should be flipped. Among remaining bin-pairs, we restricted to those for which the left and right bins were located within 50kb of the breakpoints of a single mCA call and for which the read orientations matched the copy number change called from read-depth, suggesting that the discordant reads in the bin-pair had localized the breakpoints of that mCA. Finally, for a small fraction of mCAs with multiple surviving bin-pairs (288 out of 4,520 mCAs), we chose the bin-pair with the smallest combined distance from the called breakpoints. This gave a set of 3,500 simple deletions and 1,020 tandem duplications for which one or more pairs of discordant reads localized the mCA breakpoints to approximately 1kb.

### Identifying exact mCA breakpoints using split reads

WGS data can also contain single sequencing reads composed of chimeric DNA sequence generated by an mCA, enabling the breakpoints of the mCA to be precisely determined. To find such reads, we identified “split” reads with a supplementary alignment for which the primary and supplementary alignments mapped within 2kb of a pair of breakpoints supported by discordant reads. We filtered out reads with multiple supplementary alignments and reads with combined mapping quality (summed across the primary alignment and the supplementary alignment) less than 90. We further restricted to reads with a CIGAR string matching the regular expression *^[0-S]*M[0-S]*[H,S]:* or *^[0-S]*[H,S][0-S]*M:*. Finally, we restricted to mCAs with split reads supporting exactly one breakpoint at base pair resolution (as determined from the split alignment data).

We computed the number of base pairs of microhomology between the two breakpoints of an mCA as the largest 𝑁 for which the 𝑁 rightmost aligned bases of GRCh38 for the forward-mapped read matched the 𝑁 leftmost aligned bases of GRCh38 for the reverse-mapped read.

### Calling mosaic 13q14 deletions from WGS read-depth

To increase power to detect del(13q14) mCAs, which are relatively common and usually span a specific ∼1Mb commonly deleted region on chromosome 13q starting at the *DLEU2* lncRNA, we implemented a separate detection method based on WGS read-depth in this region (chr13:49982549-50982549). For each individual, we computed a 13q14 read-depth z-score by comparing the observed number of WGS reads aligned in this region (after applying the filters used in our genome-wide read-depth profiling analysis) to the expected number of aligned reads (according to the WGS sample’s genome-wide read-depth profile, assuming no mCA in this region). We computed a provisional z-score for the observed 13q14 read count assuming a Poisson distribution with mean equal to the expected number of aligned reads (which we then approximated with a Gaussian). We then recalibrated the provisional z-scores to account for overdispersion from the Poisson distribution due to unmodeled technical influences on WGS read-depth. We performed recalibration based on the distribution of provisional z-scores observed among a subset of individuals expected to be minimally affected by del(13q14): specifically, individuals younger than 45 years of age with |𝑧_provisional_| < 4. We computed the mean and standard deviation of the provisional z-scores observed among these individuals and then recalibrated all provisional z-scores by subtracting off this mean and dividing by this standard deviation. We then called mosaic 13q14 deletions by setting a recalibrated z-score threshold of -3.5, such that individuals with recalibrated z < -3.5 were considered to have evidence of del(13q14).

We selected the threshold of z < -3.5 based on the following calculations indicating that it provides good control of false discovery rate (FDR). First, assuming the recalibrated z-scores have a standard normal distribution under the null hypothesis (no 13q14 deletion), we determined that the expected false positive rate (Φ(−3.5)=0.00023) was 4.1% of the detection rate, suggesting an FDR of 4.1%. We corroborated this estimate with an orthogonal approach that utilized the principle that clonal events associate with age. We took the mean age of 61.5 years in individuals with z < -5 as the mean age for individuals with true mosaic 13q14 deletions and the mean age of 56.2 years in individuals with z > 0 as the mean age for individuals without mosaic events. Assuming that each z-score bin contains a mixture of true positive calls with mean age 61.5 and false positive calls with mean age 56.2, we then estimated the FDR in our 13q14 calls as the proportion 𝛼 in the equation (mean age of individuals with calls) = (1 − 𝛼) ⋅ 61.5 + 𝛼 ⋅ 56.2. Using this method, we estimated an FDR of 7.4%.

### Identifying breakpoints of mosaic 13q14 deletions

We sought to localize del(13q14) breakpoints for individuals with read-depth-based evidence of mosaic 13q14 deletion to better characterize the breakpoint distribution and clonal fraction distribution of 13q14 deletions. For individuals with 13q14 read-depth z-score < -3.5, we extracted all discordant read pairs mapped to chromosome 13 with insert sizes between 5kb and 5Mb that were consistent with a deletion overlapping chr13:49982549-50982549. Individuals had on average of 3.2 discordant read pairs, often indicating different breakpoints, which posed a challenge for determining which subset of discordant read pairs corresponded to true del(13q14) breakpoints: read-depth profiles of several individuals showed clear evidence of multiple 13q14 clonal expansions with different breakpoints, as previously observed in UKB^21^. We handled these challenges by performing forward variable selection to identify a small subset of discordant read-supported breakpoint pairs (typically one or two) that could explain local deviations in read-depth measurements in the 13q14 region.

Specifically, for each discordant read pair 𝑖 = 1, …, 𝑚, we used our read-depth analysis pipeline to compute the numbers of observed reads 𝑂_𝑖_ and expected reads 𝐸_𝑖_ mapped to the region of length 𝐿_𝑖_ spanned by the discordant read pair (where 𝐸_𝑖_ was computed assuming a copy number of 2, i.e., no mCAs). A mosaic 13q14 deletion with breakpoints corresponding to the discordant read pair 𝑖 and with cell fraction 𝛽_𝑖_ would be expected to reduce read-depth in this region by 0.5 ⋅ 𝛽_𝑖_ ⋅ 𝑐, where 𝑐 denotes the sample’s WGS coverage, resulting in a depletion in read count (𝑂_𝑖_ − 𝐸_𝑖_ < 0) of magnitude 0.5 ⋅ 𝛽_𝑖_ ⋅ 𝐿_𝑖_ ⋅ 𝑐. Similarly, a mosaic 13q14 deletion with different breakpoints corresponding to a different discordant read pair 𝑗 with cell fraction 𝛽_j_ would be expected to deplete the read count in region 𝑖 by 0.5 ⋅ 𝛽_j_ ⋅ 𝐿_𝑖j_ ⋅ 𝑐, where 𝐿_𝑖j_indicates the length of overlap between regions 𝑖 and 𝑗. We took 𝑐 to be the mean coverage across 1kb bins in the 11Mb region containing the focal 1Mb deletion region and its 5Mb flanking regions.

Based on the above reasoning, we sought to identify a minimal subset of discordant read pairs 𝑆 ⊆ {1, …, 𝑚} for which a choice of mosaic cell fractions 𝛽_j_ ≥ 0 could be found such that the following set of 𝑚 equations (for 𝑖 = 1, …, 𝑚) was approximately satisfied:

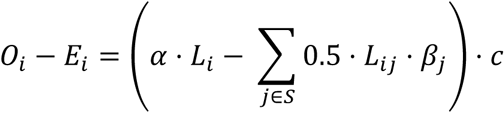

In these equations, we included an additional “intercept” term 𝛼 ⋅ 𝐿_𝑖_ ⋅ 𝑐 (where 𝛼 is a constant independent of 𝑖) to account for the possibility of slight miscalibration of the read-depth model across the 13q region.

To identify such a subset of breakpoint pairs and corresponding cell fractions 𝛽_j_, we used the following iterative forward variable selection heuristic. At each iteration, we evaluated the possibility of augmenting the current selection 𝑆 ⊂ {1, …, 𝑚} with each remaining 𝑠 ∉ 𝑆. For each such possibility, we computed the optimal choice of cell fractions 𝛽_j_ ≥ 0 (for 𝑗 ∈ 𝑆 ∪ {𝑠} using nonnegative least squares to minimize the sum of squared residuals in the 𝑚 equations above, where we normalized the squared residual of equation 𝑖 by dividing it by 𝐸_𝑖_ (to put each normalized residual on the same 𝜒^2^ scale, assuming an approximately Poisson distribution of read counts with mean and variance 𝐸_𝑖_). We then selected the augmentation 𝑠 ∉ 𝑆 that minimized the sum of normalized squared residuals. We stopped the iteration when this sum of normalized squared residuals no longer exceeded 9 (which for a single equation would correspond to the amount of unexplained read-depth deviation dropping to |z|<3).

Our approach identified at least one potential breakpoint for 1,590 out of 2,774 individuals with 13q14 depth z-score < -3.5. Among the 1,590 individuals, a total of 2,087 breakpoint pairs were identified.

### CLL survival analysis

We defined a CLL phenotype for the survival analysis based on histology and behavior codes (histology = 9823 and behavior = 3) reported in the NHS cancer registry for participants residing in England or Wales and the NHS Central Register (NHSCR) for participants residing in Scotland (1,010 cases in total). We removed 2,232 individuals with a prevalent blood cancer diagnosis (histology code >=9590 with diagnosis date preceding sample collection date). Among the remaining individuals (who constituted the “at-risk” set at the beginning of the study), we restricted analysis to individuals with European genetic ancestry. We stratified those individuals with a del(13q14) and/or 13q CN-LOH mCA by mCA type and by cell fraction, and among individuals with neither mCA, we identified an age- and sex-matched control subset by propensity score matching in a logistic regression of del(13q14) status on age and sex. We right-censored individuals with death dates reported in the NHS or NHSCR death registries. We also right-censored all cancer outcome data collected after 2018-01-01 as suggested by UKB and restricted the survival analysis to 10 years of follow-up. We used the *lifelines* package^90^ to compute Kaplan-Meier curves and confidence intervals.

In our analysis of del(13q14) progression to CLL, we removed individuals with 13q CN-LOH clones and restricted to del(13q14) calls with breakpoints that had been localized by discordant read pairs, as this allowed more accurate estimation of mCA cell fraction (by using read-depth in the entire deleted region rather than just the most focal 1Mb region).

We chose the highest cell fraction clone for individuals with multiple del(13q14) or 13q CN-LOH clones. In our analysis of individuals with 13q CN-LOH clones, we included all such individuals regardless of del(13q14) status.

### GWAS of del(13q14) and CLL

We performed genome-wide association analysis on both our WGS-derived del(13q14) + trisomy 12 phenotype and CLL diagnosis status of UKB participants, restricting to individuals with European genetic ancestry (as previously defined using principal components^91^), TOPMed-imputed genotypes, and less than 10% missingness of SNP-array genotypes. For the del(13q14) + trisomy 12 GWAS, we further restricted to individuals with available WGS data, comprising 3,470 cases (the union of 2,727 cases of del(13q14) based on read-depth z < -3.5 and 889 cases of mosaic trisomy 12) and 445,815 controls. To maximize power for the CLL GWAS, we merged all reports of the C91.1 ICD-10 code from the cancer registry (1,007 cases), hospital episode statistics (1,237 cases), and death registry (251 cases), comprising a total of 1,502 CLL cases. Restricting to individuals with European genetic ancestry, TOPMed-imputed genotypes, and less than 10% SNP-array missingness left 1,393 CLL cases and 451,346 controls. We computed association tests using regenie’s approximate Firth logistic regression^92^ controlling for sex, age, age squared, smoking status, and the first 20 genetic principal components (PCs) as covariates. We fit regenie step 1 on the SNP-array genotypes and tested TOPMed-imputed variants with MAF > 0.001 in step 2. To compare our GWAS results with results from meta-analysis of larger CLL cohorts, we downloaded the significant CLL GWAS hits in Law et al. 2017^45^ from the GWAS catalog^93^.

### Burden tests for association of rare coding variants with CN-LOH in *cis*

To define sets of protein-coding variants within genes to use for burden testing, we used variant annotations from the gnomAD v4.1.0 exomes release^94^, which included 416,555 exome-sequenced UKB participants. We extracted annotations for all protein-coding variants with high or moderate VEP consequence^95^, FILTER=PASS, and a >90% call rate in UKB WES, from which we identified missense variants and high-confidence loss of function (LoF) variants with no LoF_filter or LoF_flags; we further annotated missense variants with PrimateAI-3D scores^96^. We extracted genotypes of these variants from the UKB population-level DRAGEN WGS bgen files and dropped variants with allele frequencies discordant with frequencies reported for non-UKB non-Finnish Europeans in the gnomAD v4.1.0 data set (AF disagreement >10-fold). Finally, we merged these genotypes with LoF CNV calls we had previously generated from UKB WES data^97^.

We defined up to 64 burden masks per gene based on the following possible options for variant inclusion:

- Maximum allele frequency of 0.01, 0.001, 0.0001, or singleton (4 possible choices)
- Inclusion of LoF variants only, or inclusion of missense variants with PrimateAI-3D score >0.6, >0.7, or >0.8 (4 possible choices)
- Inclusion versus exclusion of LoF CNV calls (2 possible choices)
- Inclusion of only variants with coding consequences for the MANE_SELECT transcript^98^ versus inclusion of protein-coding variants for any transcript (2 possible choices)

In total, we constructed 1,112,952 burden masks for 17,893 genes using regenie step 2 with flags *--skip-test* and *--write-mask*, which coded individuals with a genotype of 1 if they carried at least one variant in the mask and 0 otherwise.

We tested each burden mask for association with CN-LOH mutations in *cis* using Fisher’s exact test on the binary phenotype of whether or not an individual carried a CN-LOH mutation overlapping the gene being tested. As in our previous SNP-array-based analysis^22^, we removed related individuals by preferentially keeping individuals with CN-LOH calls and older controls. We applied the Benjamini-Hochberg procedure to control the FDR at 0.01, which corresponded to p<1.2×10^-5^.

We proceeded to phase the burden mask genotypes onto our common-variant scaffold haplotypes to determine whether CN-LOH mutations that associated with rare coding variants in certain genes tended to make these variants homozygous or remove them from the genome. We performed both statistical phasing using SHAPEIT5 *phase_rare* (filtering to confident phase calls, i.e., phase probability PP>0.8) as well as read-based phasing using a custom pipeline to identify reads or read pairs that overlapped both a rare coding variant and a nearby common variant, thereby enabling the rare coding variant to be phased onto the common-variant scaffold. Specifically, we first used samtools mpileup (−Q 20) to find phase-informative reads (or read pairs) within a 1kb of a target variant to be phased. For each nearby heterozygous variant, we counted the number of fragments (reads or read pairs) supporting the variant having the same phase vs. opposite phase relative to the target variant. We restricted to nearby variants for which phase-informative fragments unanimously supported either the same or opposite phasing of REF alleles. We then merged the resulting phase sets into our common-variant scaffold to determine phase relative to CN-LOH, dropping phase sets containing variants whose relative phase disagreed with the scaffold phase.

For each of the 38 genes with FDR-significant burden associations, we evaluated the directionality of CN-LOH mutations that were confidently phased relative to rare coding variants by the above pipeline. We used a Bayesian approach to decide whether the observed fraction of CN-LOH mutations that made rare coding variants homozygous reflected a confident skew away from 0.5. Specifically, we computed the Bayesian two-sided tail probability 2 ⋅ min (𝑃(𝑎 < 0.5 ∣ phase counts), 𝑃(𝑎 > 0.5 ∣ phase counts)), where 𝑎 is the fraction of CN-LOH mutations that make rare coding variants in the gene homozygous, for which we assume an uninformative 𝐵𝑒𝑡𝑎(0.5, 0.5) Jeffreys prior, and posterior probabilities of 𝑎 < 0.5 and 𝑎 > 0.5 are computed given the observed counts of CN-LOH mutations with each phase relative to rare coding variants. We considered the gene to have a confident skew in CN-LOH directionality if this two-sided tail probability was less than 0.1.

To perform gene ontology (GO) enrichment analysis on the 38 genes identified by the burden tests, we used *gseapy*^99^ to test for enrichments of these genes in the GO_Biological_Process_2025^100^, KEGG_2021_Human^101^, and Reactome_Pathways_2024^102^ gene sets.

For our analyses of CN-LOH directionality beyond the 38 genes with FDR-significant burden associations, we defined DepMap.Q10 genes as the 480 genes in the lowest decile of average DepMap scores^103^ across all lymphoid and myeloid cell lines. For each such gene, we identified carriers of rare (AF<0.001) damaging variants (LoF or missense with PrimateAI-3D score > 0.8 in any transcript) for which the variant could be confidently phased relative to an overlapping CN-LOH mutation. We then determined whether the majority of phased damaging variants in the gene under consideration were made homozygous or were removed by CN-LOH mutations, dropping genes for which these counts were tied; this left 379 genes with nonzero, non-tied counts. Most counts were small, such that at the level of individual genes, we expected the majority direction of CN-LOH mutations to be a noisy indication of whether damaging variants were pro- or hypo-proliferative, but across hundreds of genes, a pattern could be observed. Similarly, we annotated genes as tumor suppressor genes and oncogenes based on the MSK OncoKB database^78,104^ and then restricted analyses to genes with one or more carriers of damaging variants with overlapping CN-LOH mutations as above.

### Common-variant GWAS for CN-LOH in *cis*

We also tested the common variants (MAF > 0.01) that we had imputed from the SHAPEIT5 200K reference panel for association with CN-LOH mutations in *cis*. We coded heterozygous genotypes as 1 and homozygous genotypes as 0 (under the model that a difference in proliferative potential of the two alleles should be observed in heterozygotes but unobserved in both hom-REF and hom-ALT individuals) and used Fisher’s exact test with the binary phenotype of whether or not an individual carried a CN-LOH mutation overlapping the variant being tested. We restricted analysis to unrelated individuals with European genetic ancestry, pruning related individuals as in the burden analysis. We further removed individuals with CN-LOH mutations potentially explained by rare (AF<0.01) coding variants (including CNVs) in the 38 genes with FDR-significant burden associations (i.e., we removed individuals with a CN-LOH mutation that overlapped an LoF variant or a missense variant with PrimateAI-3D score >0.6 in any transcript of one of the 38 genes); this mitigated the tendency for some common variants to tag associations driven by haplotypes containing large-effect rare variants. Finally, we also removed CN-LOH mutations that appeared likely to be driven by two rare splice region variants in *MPL* and *ATM*: chr1:43338725:G:C (Pangolin score of 0.68) and chr11:108257471:T:G (Pangolin score of 0.82)^105^.

In addition to the above test for association between common variants and presence of a CN-LOH in *cis*, we also tested the phase of common variants for association with the directionality of overlapping CN-LOH mutations (i.e., whether or not CN-LOH mutations tended to make the alternate allele homozygous or remove it from the genome). We used a binomial test for deviation of the balance of directionality from 0.5.

### Polygenic effects of common variants associated with blood cell counts

To search for evidence of polygenic effects of common variants influencing CN-LOH, we leveraged common-variant effects on blood count phenotypes, which served as a proxy for proliferative potential conferred by common variants. First, we assessed the extent to which polygenic scores for blood count phenotypes, computed across the alleles on each homolog of a chromosome affected by a CN-LOH mutation, were informative of the directionality of the CN-LOH mutation. We followed the same approach we previously used to analyze SNP-array-based CN-LOH calls^22^, using the same polygenic scores for blood count phenotypes that we had previously computed on UKB using BOLT-LMM^106^. We restricted our analyses to individuals with European genetic ancestry and removed individuals with CN-LOH potentially explained by rare coding variants in the 38 genes with FDR-significant burden associations (as in the common-variant GWAS).

Second, we analyzed specific variants associated with white blood count (WBC) for potential effects on CN-LOH directionality. We analyzed GWAS summary statistics for WBC previously computed on UKB using BOLT-LMM^107^. For each common WBC-associated index variant (MAF>0.01, clumped within a 1Mb window), we determined whether heterozygous carriers of the variant who experienced a CN-LOH mutation had the WBC-increasing allele amplified more often than removed; if so, we considered the variant to have CN-LOH direction consistent with the effect direction of the WBC-associated variant. We performed the analysis at various cutoffs for minimum absolute effect size on WBC.

### Heritability of CN-LOH directionality

To estimate the extent to which common variants on each chromosome arm collectively influence the directions of CN-LOH mutations on that arm, we performed a heritability analysis under a liability threshold model. Our model assumes that each observed CN-LOH clonal expansion is driven by a differential underlying proliferative potential of a cell with the CN-LOH mutation vs. other cells, 𝑦_𝑝𝑟𝑜𝑙𝑖𝑓_, which we model as

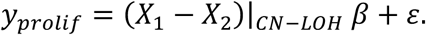

Here, (𝑋_1_ − 𝑋_2_)|_𝐶𝑁–𝐿0𝐻_ is the difference between normalized minor allele counts on the two haplotypes (coded as 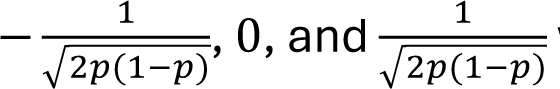 where 𝑝 indicates allele frequency) restricted to common variants within the boundaries of the CN-LOH mutation (and equal to zero elsewhere), 𝛽 is a vector of variant effect sizes for proliferation, and 𝜀 encapsulates all other effects on proliferation (e.g., somatic mutations that confer proliferative potential to the cell distinct from other cells). The observed direction of the CN-LOH then depends on the sign of 𝑦_𝑝𝑟𝑜𝑙𝑖𝑓_: when 𝑦_𝑝𝑟𝑜𝑙𝑖𝑓_ > 0, CN-LOH mutations that replace alleles on haplotype 2 with those on haplotype 1 have a proliferative advantage and expand, and when 𝑦_𝑝𝑟𝑜𝑙𝑖𝑓_ < 0, the opposite is true (under a parallel scenario in which the sign of 𝜖 is flipped). The common-variant heritability of the differential proliferative potential that determines CN-LOH direction is then

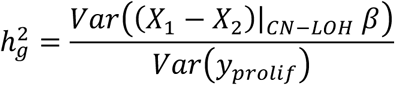

where the variances take into account randomness in the span of CN-LOH mutations that span different genomic intervals. We can estimate ℎ^2^ under the standard liability threshold model using restricted maximum likelihood (REML, as implemented in GCTA^64^) by randomly assigning haplotypes to 𝑋_1_ and 𝑋_2_ such that the “case prevalence” is 0.5. We verified in simulations that REML produces unbiased estimates of ℎ^2^ across a wide range of parameter settings under this model of CN-LOH directionality (**Supplementary Fig. 7**).

We applied this model to estimate the common-variant heritability of CN-LOH directionality for 31 chromosome arms with at least 200 CN-LOH carriers (after excluding CN-LOH mutations containing rare coding variants in the 38 putative target genes). We performed LD-pruning on the common variants that we imputed for mCA detection, excluding variants with a greater than 5% missingness, using plink2^108^ with a 500kb window and r2 cutoff of 0.5. For each chromosome arm, we dropped CN-LOH events that overlapped the other arm of the chromosome (thereby dropping isodisomy events) and computed a “kinship matrix” using the difference in haplotypes within CN-LOH boundaries after correcting switch errors identified during mCA calling. We normalized the “kinship matrix” by the mean of its diagonal to account for differences in lengths of various CN-LOH events and supplied the resulting covariance matrix to GCTA^64^ with the *--grm* flag and the *--reml*, *--reml-no-constrain*, and *--prevalence* 0.5 flags to fit our liability threshold model with unconstrained REML. We reported the liability-scale ℎ^2^ estimates from GCTA as our estimates for the heritability of CN-LOH directionality.

