## Supplementary Figures for "Patterns and drivers of 43,617 mosaic chromosomal alterations in blood"

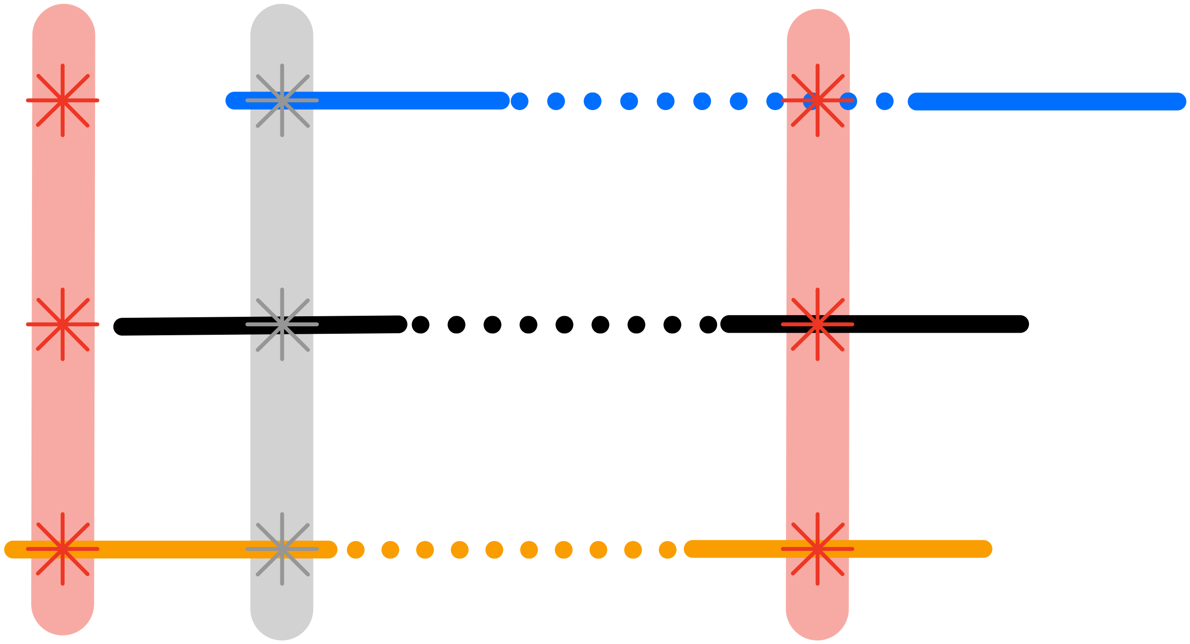

**Supplementary Figure 1: Counting sequenced DNA fragments that overlap heterozygous sites.** Paired-end reads derived from a DNA fragment that overlaps one or more of an individual's heterozygous sites can be assigned to its haplotype of origin, such that the allelic balance between an individual's two haplotypes can be evaluated by counting and comparing the numbers of fragments attributable to the two haplotypes. However, it is not possible to precisely count such fragments using only variant-level information about allelic depths (the AD field in VCF files). The reason is that a single DNA fragment (which generates a pair of sequencing reads) may contribute to allelic depth measurements at multiple nearby heterozygous variants (indicated with stars in the schematic above), such that summing allelic depths across heterozygous variants double-counts some fragments. An approximate way to ameliorate this issue (implemented by MoChA) is to "thin" the set of heterozygous variants under consideration: i.e., to exclude allelic depths measured at variants that are too close to another nearby variant (e.g., the gray variant in the schematic). This heuristic approach works reasonably well but is not optimal: in the schematic above, it correctly counts the black fragment, double-counts the orange fragment, and drops the blue fragment. To instead count each fragment exactly once, it is necessary to analyze read-level data from sequence alignments (bam/cram files).

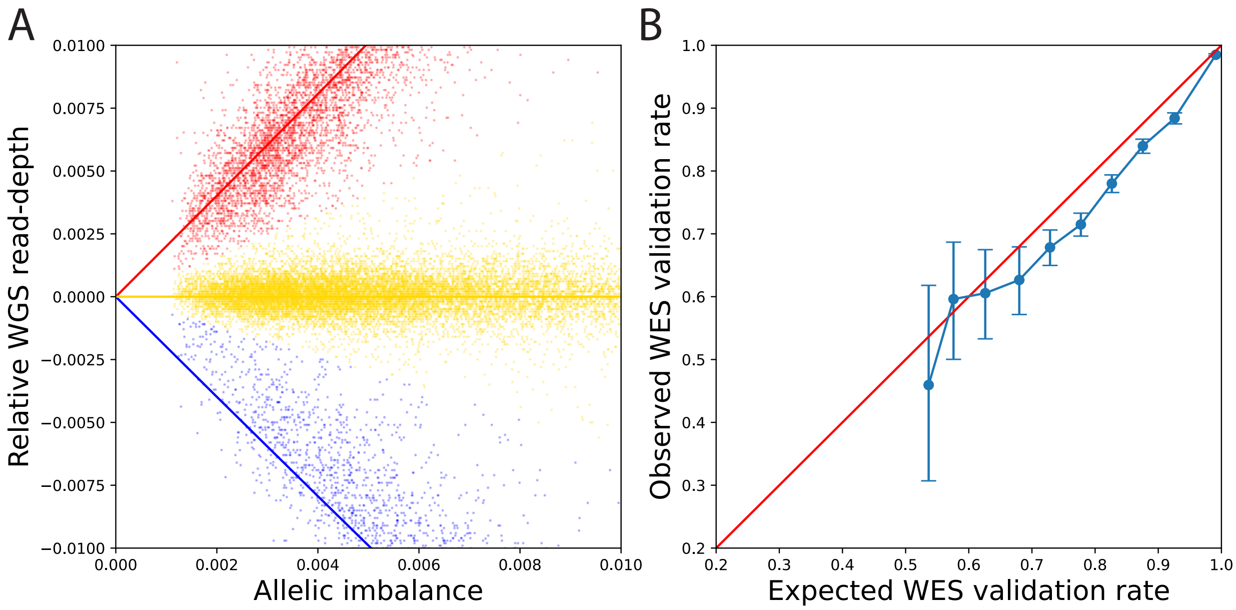


**Supplementary Figure 2: Copy-number state determination and WES-based validation of mCAs. (A)** Zoom-in of **Fig. 1c**: WGS read-depth deviation (i.e., fractional increase or decrease relative to expectation assuming diploid copy number) and allelic imbalance within the boundaries of mCAs cluster along three curves corresponding to the relationship between these quantities for gains (red), CN-LOH mutations (gold), and losses (blue). Even among mCAs with the lowest levels of allelic imbalance, most mCAs had WGS read-depth measurements close enough to one of the three expected values to confidently classify copy-number state. A very small fraction of mCAs had intermediate read-depth values and ambiguous copy-number state; because these mCAs represented a negligible fraction of the call set, we generated a best-guess copy-number state for all mCAs (based on linear separators; **Methods**) rather than leaving some mCAs with undetermined copy-number state. **(B)** mCAs called from WGS data exhibit consistent allelic imbalance of exome sequencing reads. The validation rate is computed as the fraction of mCAs with concordant directionality of WGS and WES allelic imbalance. Expected validation rates were computed at a per-mCA level based on the estimated mCA cell fraction and WES coverage within the mCA (**Methods**); observed validation rates are shown for mCAs stratified into tranches of expected validation probability. Error bars, 95% Jeffreys intervals.

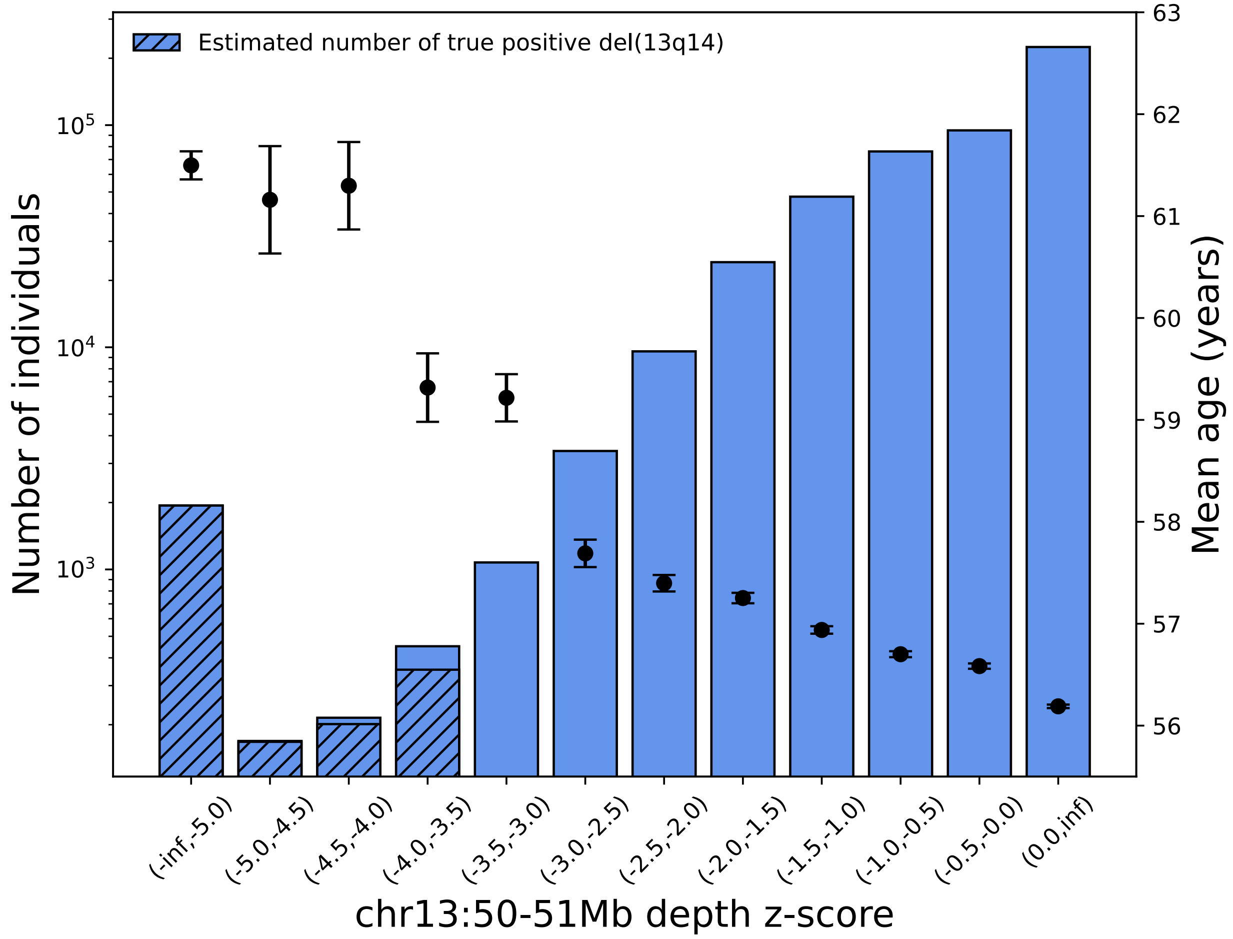


**Supplementary Figure 3: Estimated FDR for del(13q14) calls derived from WGS read-depth.** The FDR for mosaic 13q14 deletion calls was first estimated theoretically (diagonal hatching) by comparing the null vs. observed distribution of WGS read-depth z-scores and then validated using the age distribution of individuals within z-score bins (the intuition being that true positive 13q14 deletions are expected to be enriched in older individuals while false positive calls are expected to show no association with age; **Methods**). Setting the 13q14 deletion threshold at z<-3.5 controls the FDR at approximately 4.1% based on enrichment of z-scores and 7.4% based on enrichment of older individuals.

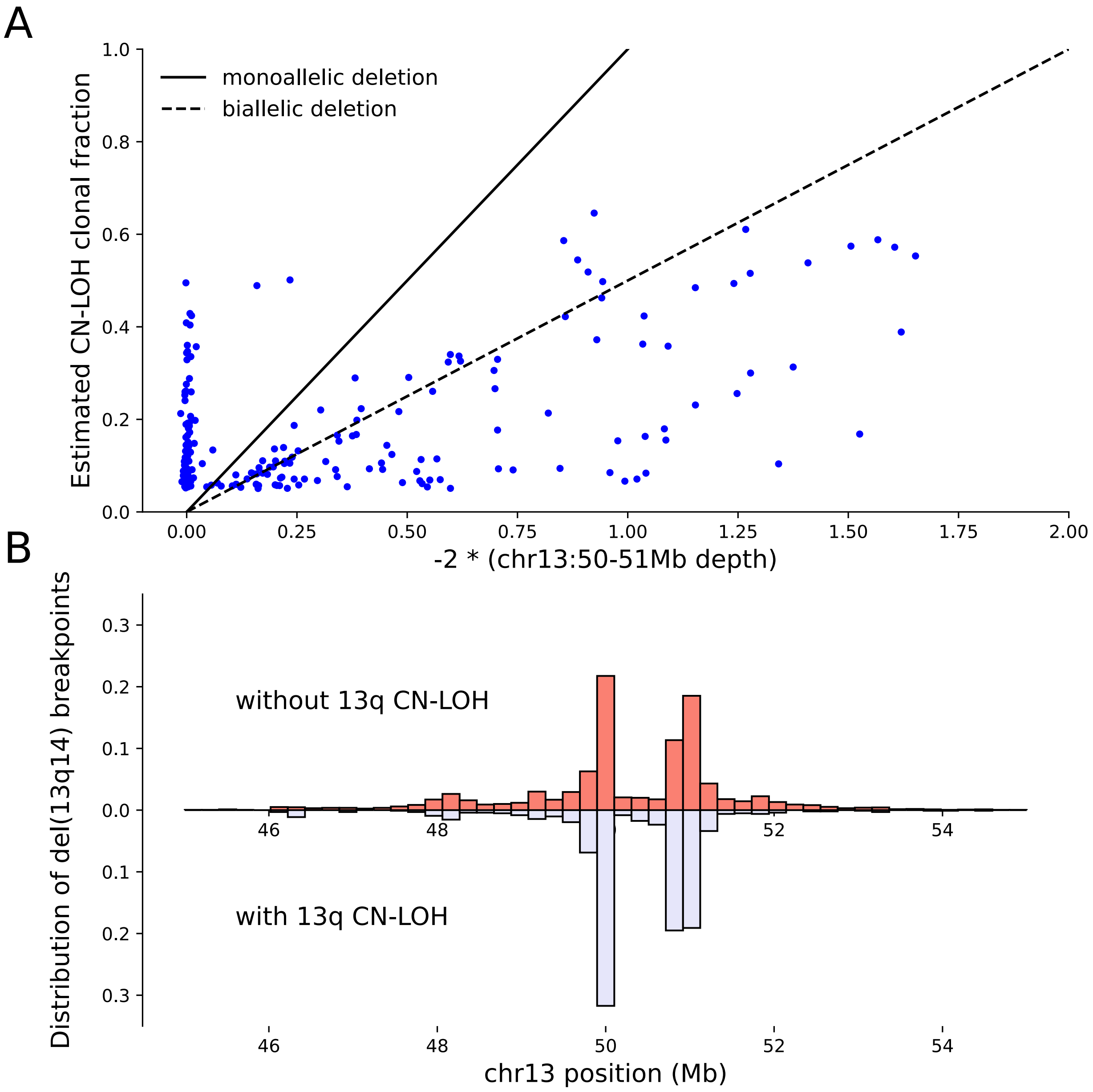


**Supplementary Figure 4: Most but not all mosaic 13q CN-LOH mutations appear to generate biallelic deletion of 13q14. (A)**119 of 199 individuals with 13q CN-LOH called at cell fractions greater than 0.05 have WGS read-depth within chr13:49982549-50982549 consistent with the CN-LOH mutation creating a “second hit” biallelic deletion of 13q14 in the expanded clone. As in other figures, WGS read-depth deviation was computed as the fractional increase or decrease relative to expectation assuming diploid copy number (such that observing no reads in the region would correspond to a depth deviation of –1). **(B)** Individuals with both 13q14 deletions and 13q CN-LOH mutations tend to have shorter 13q14 deletions, suggesting selective pressure in flanking regions preventing longer deletions from becoming biallelic.

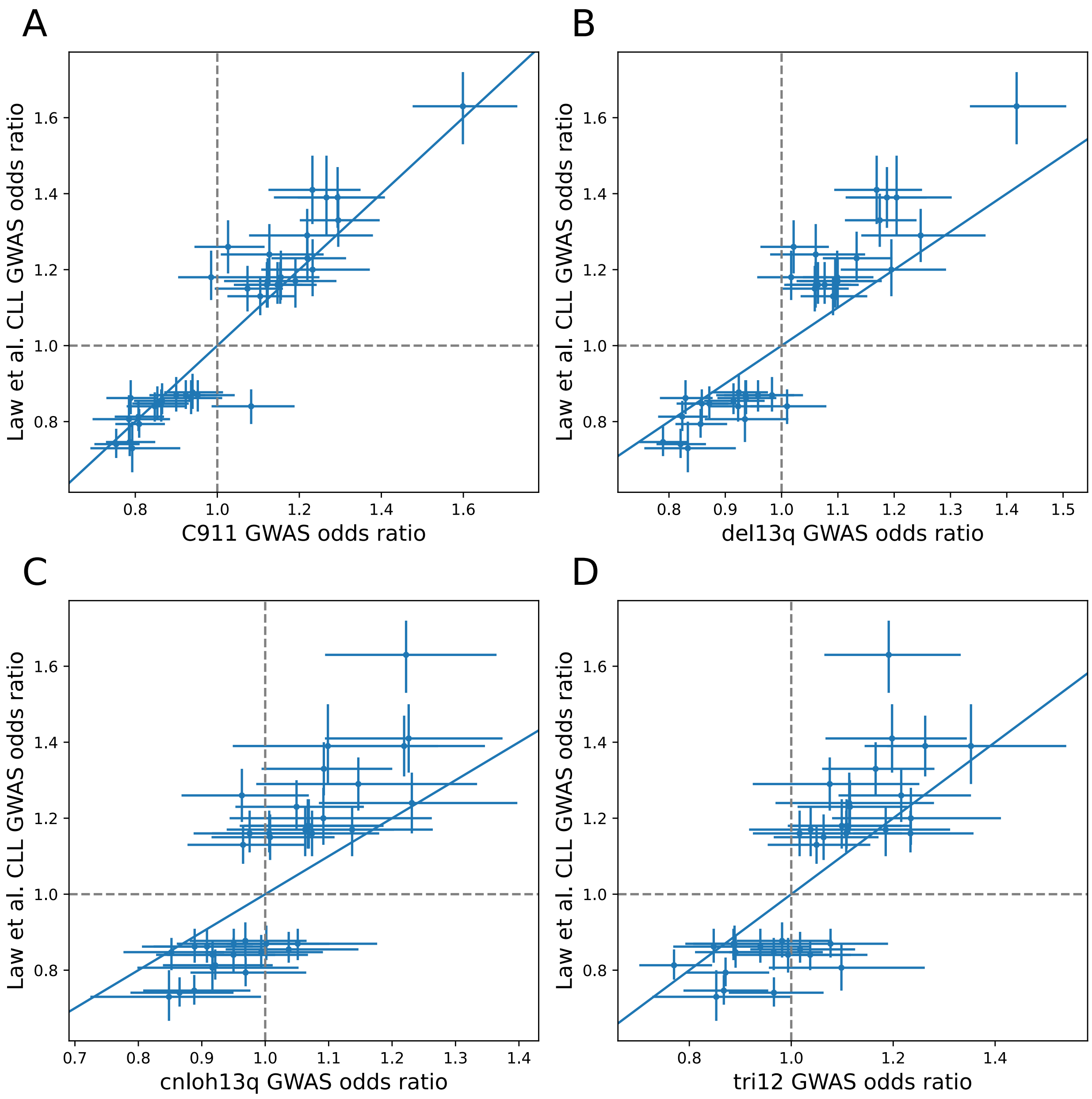


**Supplementary Figure 5: Similarity of genetic influences on CLL and CLL-associated mCAs.** Scatter plots of effect sizes of CLL-associated index variants reported by Law et al. 2017 versus effect sizes on **(A)** CLL cases in UKB, **(B)** mosaic 13q14 deletions, **(C)** mosaic 13q CN-LOH, **(D)** and mosaic trisomy 12. Error bars, 95% CIs.

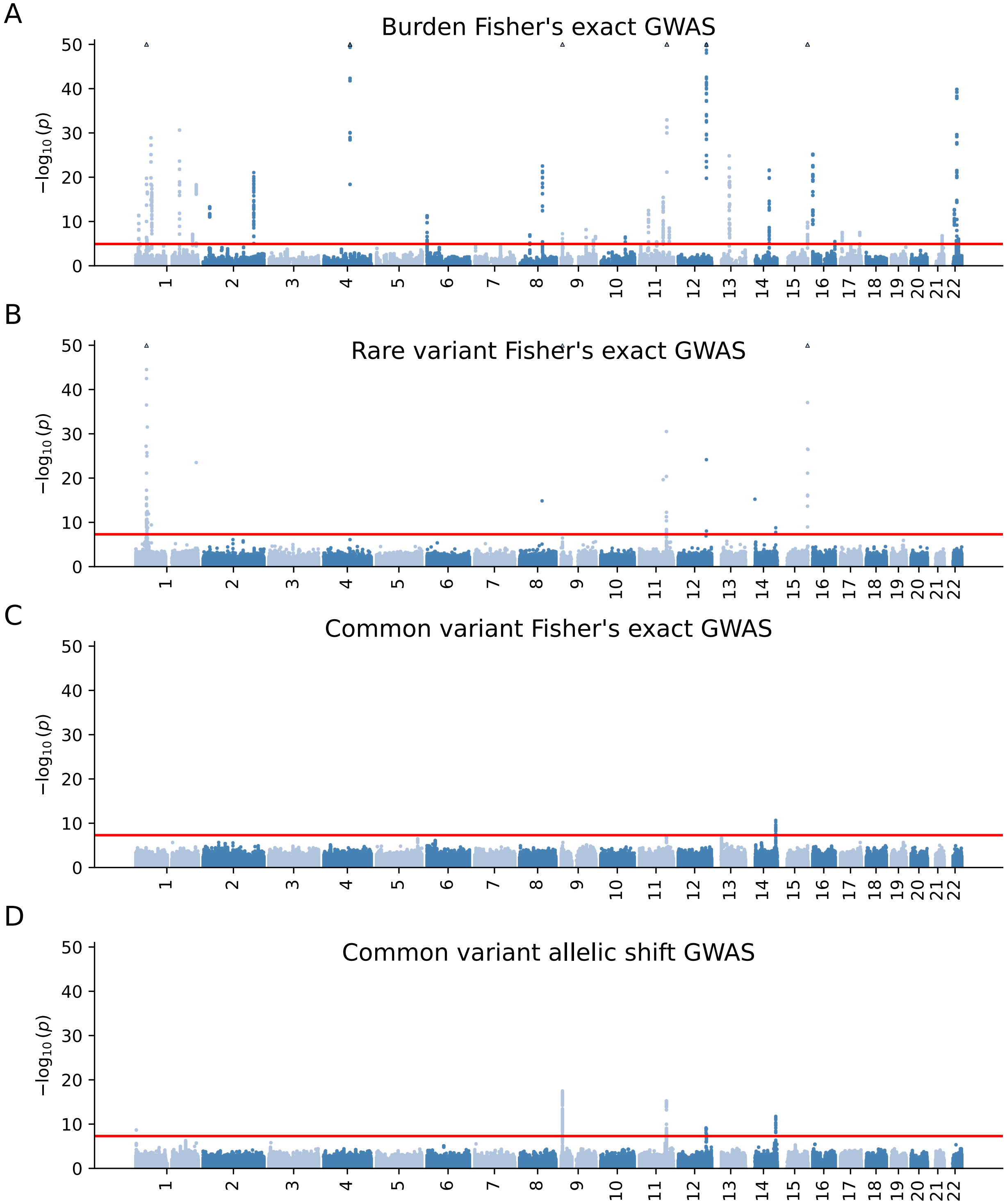


**Supplementary Figure 6: Associations of variants of various classes with the presence of an overlapping CN-LOH mutation.** Manhattan plots are provided from GWAS of *cis* CN-LOH mutations with **(A)** burden “variants” using Fisher’s exact test, **(B)** individual rare protein-coding variants (MAC>=20) using Fisher’s exact test, **(C)** common variants (MAF >

0.01) using Fisher's exact test, and **(D)** common variants using the binomial test for association of alleles with CN-LOH directionality (**Methods**). Red lines indicate significance thresholds ($p<1.2\text{x}10^{-5}$ corresponding to FDR<0.01 for the burden GWAS; $p<5\text{x}10^{-8}$ for the other three GWAS).

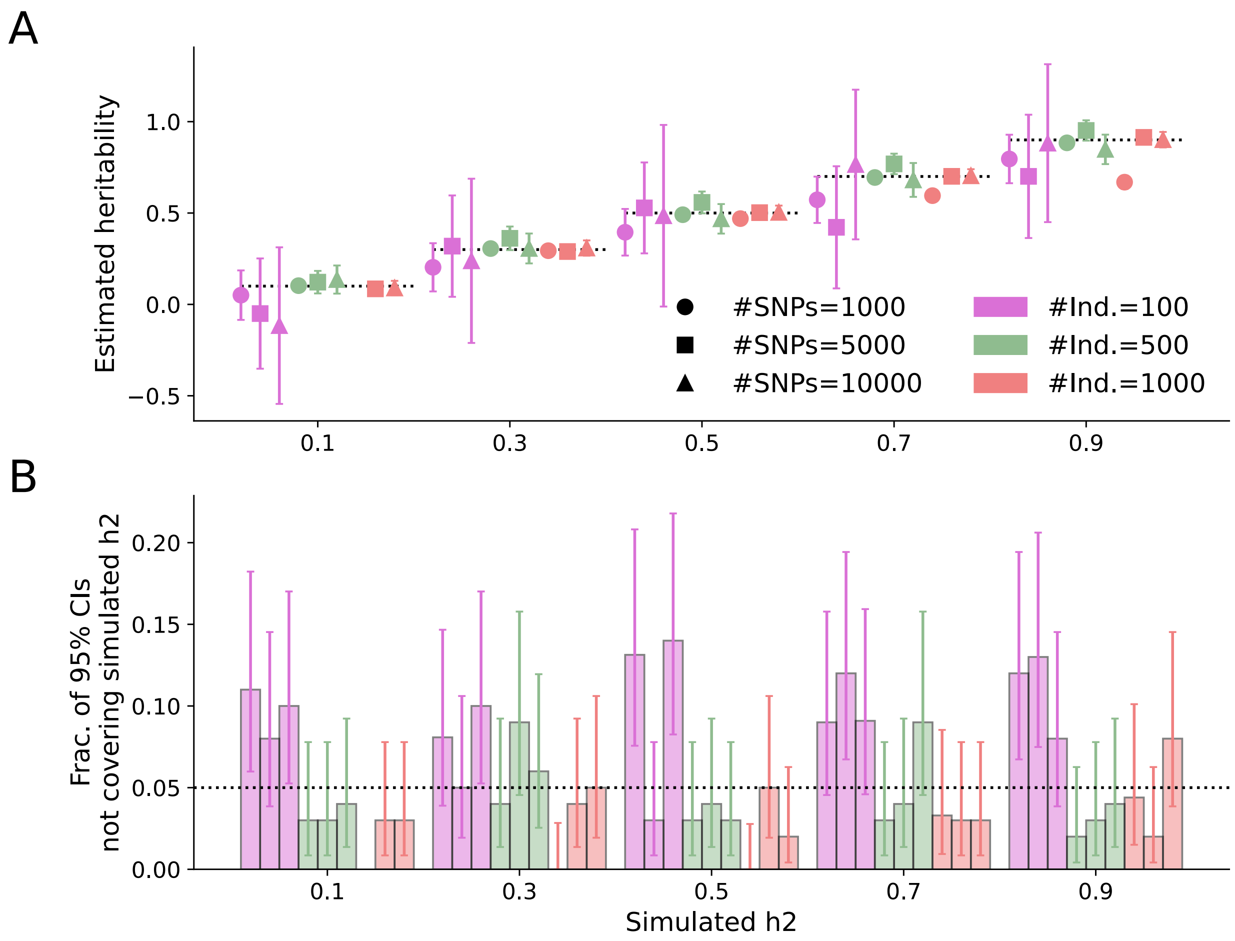


**Supplementary Figure 7: Simulations validating heritability analyses to estimate the contribution of common inherited variants to CN-LOH directionality.** Simulations under the assumed generative model of CN-LOH directionality (**Methods**) show that variance components analysis (REML) produces unbiased estimates of heritability **(A)** and calibrated confidence intervals **(B)** across a range of genetic architectures and sample sizes. We simulated genotypes using allele frequencies drawn from a uniform distribution and simulated effect sizes for all variants under an infinitesimal model of normally distributed standardized effects. For each individual, we simulated a CN-LOH event with a start position drawn uniformly from the list of variants and an end position that extended to the last variant. We zeroed out genotypes outside of the CN-LOH boundaries and computed the difference in haplotype proliferation scores within CN-LOH boundaries defined by the simulated effect sizes. We rescaled the differential proliferation scores and added normally distributed noise to simulate liability scores at a desired level of heritability. Finally, we binarized the liability scores according to their sign to obtain a CN-LOH directionality phenotype.
